# Responsive caregiving, opportunities for early learning, and children’s safety and security during COVID-19: A rapid review

**DOI:** 10.1101/2021.02.10.21251507

**Authors:** Kerrie Proulx, Rachel Lenzi-Weisbecker, Rachel Hatch, Kristy Hackett, Carina Omoeva, Vanessa Cavallera, Bernadette Daelmans, Tarun Dua

## Abstract

**Introduction:** During the COVID-19 pandemic, there have been drastic changes in family life and programs and services that promote and protect early childhood development. Global stakeholders have raised concerns that the pandemic is putting enormous strain on parents and other caregivers, compromising capabilities and enabling environments for nurturing care of young children and therefore likely impacting children’s development.

**Methodology:** This rapid review takes stock of emerging research on nurturing care for young children during the COVID-19 crisis. Two databases were searched in addition to an extensive search for grey literature, drawing on 112 scholarly and scientific studies from more than 30 countries that have examined components of nurturing care during the pandemic, namely: responsive caregiving, early learning and play, and children’s safety and security.

**Results:** There are some reports of unexpected positive benefits of the pandemic on families, including increased father involvement in caregiving. But more commonly, the studies’ findings reveal numerous issues of concern, including parental and caregiver mental health difficulties and less responsive parent-child relationships, increased screen time among children, limited opportunities for outdoor play, and fractured systems for responding to potential child neglect and maltreatment. Evidence suggests limited access and challenges in the provision of remote learning for the youngest learners, such as those in early childhood education.

**Conclusion:** The findings can inform global stakeholders, who have advocated for increased support and funding to ensure young children and other caregivers are supported and protected during the COVID-19 pandemic. There is an urgent need for action-oriented implementation studies – those that go beyond identifying trends and begin to pinpoint “what works” to effectively promote and protect nurturing care during emergencies such as the COVID-19 pandemic.

**Key questions:** *What is already known?:* The most fundamental promotive experiences in the early years of life to reach optimal development come from nurturing care and protection received from parents, family, and community, which have lifelong benefits including improved health and wellbeing. Health and other emergencies are detrimental to the provision of nurturing care.

*What are the new findings?:* Findings from this rapid review reveal numerous areas of concern, including families reporting mental health difficulties and less responsive parent-child relationships, increased screen time among children, limited opportunities for outdoor play, and fractured systems for responding to potential child neglect and maltreatment. As with other features of this pandemic, not all families are affected equally: financially vulnerable families are much more likely to experience negative ramifications. The pandemic is also disproportionately affecting parents and other caregivers with young children, particularly mothers, those with pre-existing mental health difficulties, and those caring for children with disabilities.

*What do the new findings imply?:* Findings highlight the need for action by governments, civil society, international and community-based organizations to improve support for families so that the pandemic does not break the provision of nurturing care and wipe out decades of progress, especially for vulnerable families and children.

## INTRODUCTION

During the COVID-19 pandemic, there have been drastic changes in family life on every continent. There have been prolonged containment measures in many countries, with second waves and renewed lockdowns. Families are “locked in” together, without access to their usual forms of social support and with disruptions to services and programmes that protect and promote early childhood development (i.e., childcare, early childhood education, parenting programmes, health services). Global stakeholders have raised concerns that essential components of ‘nurturing care’ for early childhood development have been harmed during the pandemic [1, 2]. The World Health Organization (WHO), UNICEF, and World Bank Group advocate that young children require five inter-related components of nurturing care for optimal development and well-being: good health, adequate nutrition, a family environment rich in responsive caregiving, opportunities for formal and informal early learning, and protection from safety risks and maltreatment [3]. During the first years of life, parents, intimate family members and caregivers are the closest to the young child and thus the best providers of nurturing care. This is why secure family environments are important for young children. In order to provide caregivers with time and resources to provide nurturing care, multisectoral policies, services and community supports need to be in place. Health, nutrition, education, social welfare, and child protection all have a role to play in creating an enabling environment for nurturing care (for further details, see https://nurturing-care.org).

Evidence shows that epidemics and pandemics are associated with disruptions to nurturing care and child development, stemming from increased caregiver social isolation and stress [4], disruption to family routines [5], traumatic stress among caregivers [6], household conflict and anxiety [7], early termination of breastfeeding [8], and reduced play among children and engagement in team-based sports [9]. This rapid review aimed to capture the emerging evidence on the effects of the COVID-19 pandemic on three components of nurturing care: responsive caregiving, early learning opportunities, and child safety and protection from violence and neglect. This is a broad inquiry that is explored through different outcomes that are critical for nurturing care and child development. For responsive caregiving, it includes good mental health of parents and caregivers, warm and responsive parent-child relationships, breastfeeding support (to promote emotional bonds and attachment), and fathers’ engagement in caregiving. For early learning, it includes access to formal and home-based learning, parent–child reading and other early learning activities, and safe play spaces. For children’s safety and security, it includes protecting children against injury, neglect and abuse, including child protection and referral services for suspected maltreatment. This review did not examine child health and nutrition, the other components of nurturing care, due to ongoing work and recently published reviews in these areas. The review findings can help identify and conceptualize priority issues for early years’ policy and programming during emergencies such as the COVID-19 pandemic and identify the literature gaps where future studies are required.

## METHODS

To generate evidence in a short time, we employed abbreviated systematic review methods. A single reviewer completed the study selection and data extraction with verification by a second reviewer, and we omitted quality assessments of the included studies. The population of interest is young children (birth to 8 years old) and their caregivers, including birth parents, adoptive parents, and formal caregivers. Studies involving children up to 18 years old and their caregivers are included as long as the studies also included results for children under 8 years old. Published quantitative or qualitative studies that document their full methodology are included, and preprints and the grey literature to ensure representativeness and due to the rapidly evolving nature of COVID-19 pandemic impact evidence.

We focused on studies that reported on responsive caregiving during the COVID-19 pandemic (e.g., parental mental health, caregiver stress, parent–child interactions, family functioning, fathers’ engagement in caregiving, gender roles and norms, breastfeeding, skin-to-skin contact, social support). We also included studies that reported on opportunities for early learning during the pandemic (e.g., access to formal and home-based learning, parent–child reading and other early learning activities, outdoor play, physical activity, screen time, sedentary behaviour). Lastly, we included studies that reported on children’s safety and security during the pandemic (e.g., child injuries, child maltreatment referrals, child physical or psychological abuse, family violence). We searched two databases (PubMed and ERIC) based on a search strategy developed and pilot-tested in collaboration with a librarian. Also, for the grey literature, the review team manually searched the websites of more than 80 multilateral and bilateral organizations, internal NGOs, foundations, and so forth. Lastly, we screened the reference lists of the included studies for further relevant citations. We restricted the search to papers available in English, French, Spanish, and Portuguese. Studies were published between 1 January 2020 and 25 October 2020.

Identified records were exported and managed on Zotero. After removing duplicates, the titles and abstracts of the remaining records were screened against the eligibility criteria, followed by full-text screening. We extracted data from the studies using a standardized form, with a second reviewer assessing for completeness and accuracy. Data extracted included, but was not limited to, investigators/authors, year of publication, setting, context, details of the methods and data collection, participants, outcomes, results, and conclusions. Data extraction was guided by Preferred Reporting Items for Systematic reviews and Meta-Analyses (PRISMA) checklist items. The preliminary stages of the synthesis involved organizing the extracted data through itemizing the data into thematic areas and tabulating the results to systematically identify patterns within and between studies and variations across studies. Because of the heterogeneity of the available primary studies, we synthesized the results narratively in the report.

## RESULTS

After excluding duplicates, the initial search yielded 4210 papers from the database search and 86 publications through the organizational website search. After the title, abstract and full-text review, 112 publications were included in the review. The PRISMA flow diagram in Figure 1 provides an overview of the study selection process.

**Figure 1.**
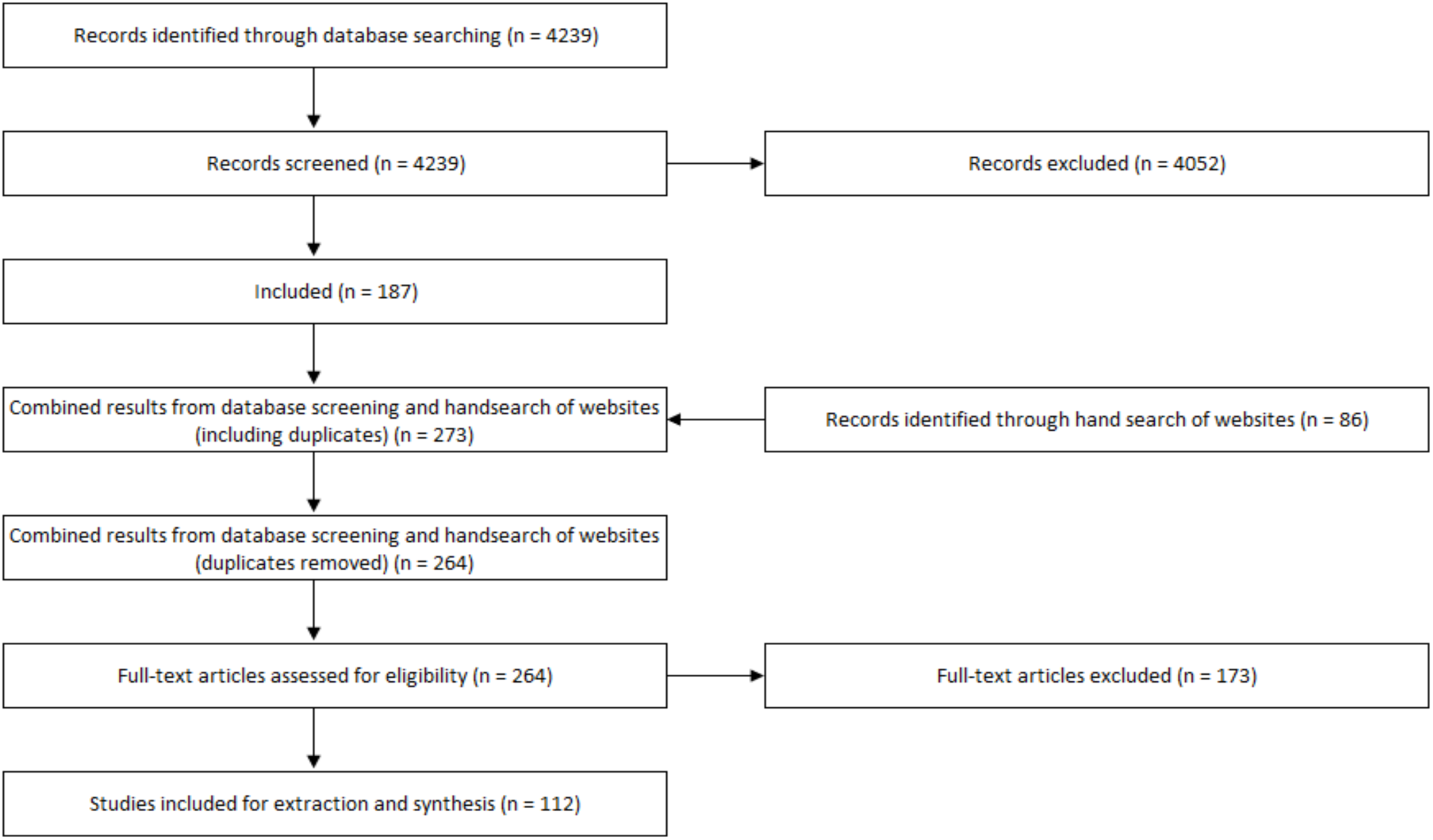
PRISMA diagram

While most studies (89) in the review focus on high-income countries, there is increasing geographic diversity from low- and middle-income countries (23). In terms of the thematic areas, the largest number of papers focused predominantly on issues related to ‘responsive caregiving’ (59), which points towards increased parent and other caregiver stress, burnout, depression and anxiety during the COVID-19 pandemic, and harsher parenting and less warm or responsive parenting. Evidence suggests increased fathers’ engagement during the COVID-19 pandemic. On the other hand, evidence indicates that COVID-19 restrictions have hampered access to traditional lactation support services. The next largest number of studies examined issues related to ‘safety and security’ (35), which point towards drop in child abuse and neglect referrals to child protection services, and a reduction in emergency department visits for child injuries. Lastly, a relatively small number of studies addressed opportunities for early learning and play during the COVID-19 pandemic (18). These studies point towards increased screen time among children, and a reduction in outdoor play and physical activities. Evidence is limited on home-based learning for young children (age 3-5 years) during the COVID-19 pandemic. What is available suggests challenges, frustrations and inequitable access to distance learning for the youngest learners. Results include 95 quantitative studies (primarily cross-sectional due to the nature of the pandemic), nine qualitative studies, and eight mixed methods designs. Key findings of included studies are synthesized in the following sections, and a summary of each study can be found in the complementary table.

**Figure 1.**
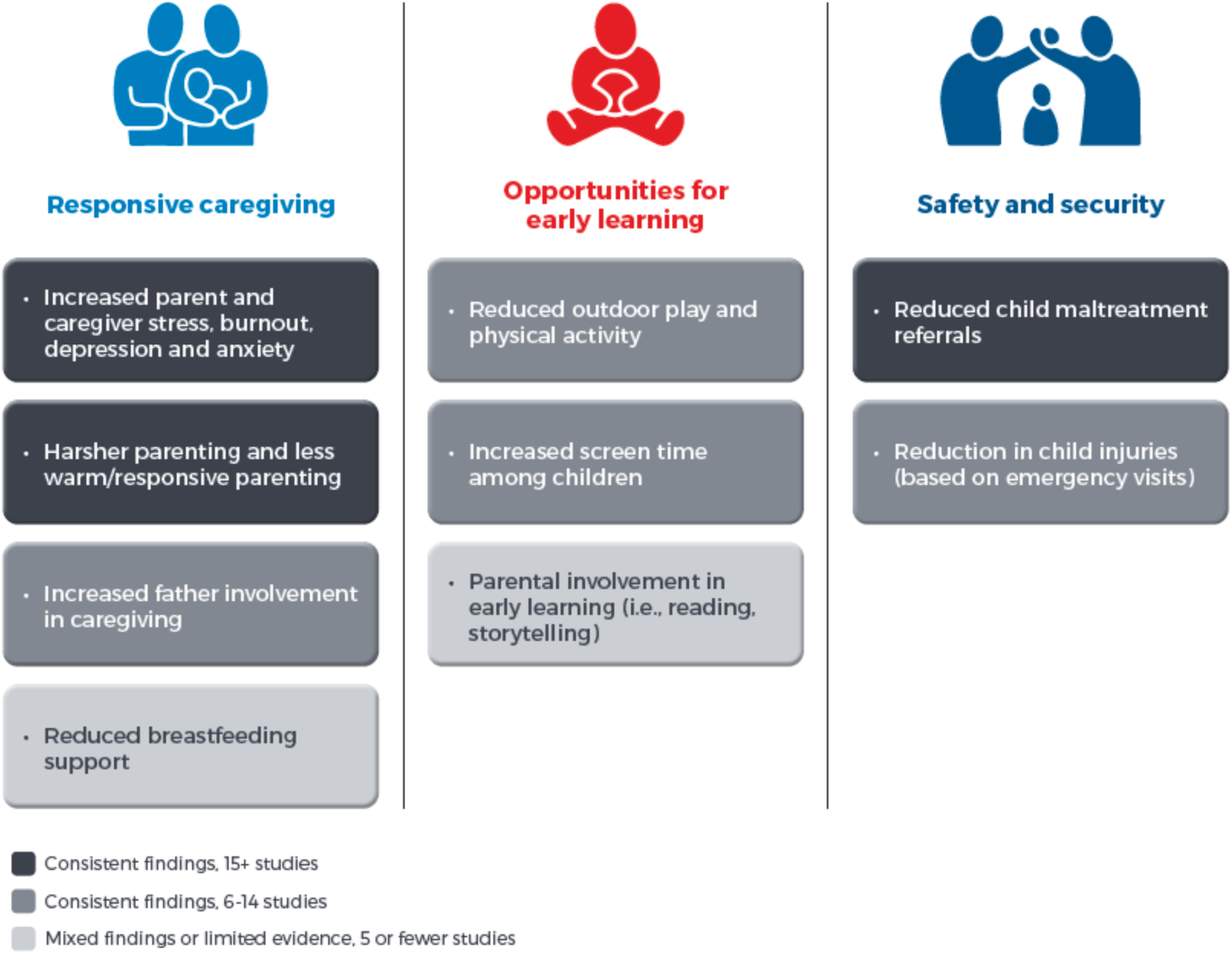
Key review findings on nurturing care during the COVID-19 pandemic.

### Parent and caregiver stress, mental health, and responsive caregiving

The review results indicate that parents’ and other caregivers’ mental health has been among the most researched topics related to nurturing care during the pandemic. During the period under review, more than 30 studies across various geographic and demographic contexts examined the implications of the COVID-19 pandemic on parent and caregiver stress, burnout, depression, and anxiety [10–45]. The presented evidence indicates that the pandemic crisis has been detrimental to caregiver stress and mental health, particularly for parents and other caregivers of young children and mothers [14,16,30,31,33], being single [30], economically vulnerable families [14,27,42], and those with pre-existing mental health issues [14, 42]. Parents and other caregivers of children with disabilities also showed a heightened prevalence of depressive symptoms, anxiety, and stress during the pandemic [11,18,43].

While this evidence base is heterogeneous, with variations in the data, context, and methodology, the findings are generally consistent in revealing that the pandemic and its related policy response measures drive increases in caregiver stress and poor mental health. For example, a study of 192 Italian mothers found that 29% of mothers who had given birth during the COVID-19 lockdown had an Edinburgh Postnatal Depression Scale (EPDS) global score above the cut-off score of >12, compared to only 12% in the control group who had given birth during the same period the year before COVID-19 [45]. A study of 651 Finnish parents of children (aged 5 to 8 years old) found that 26% met the clinical EPDS cut-off score for potential depression during the pandemic, compared to 15% among the same sample parents with data collected between 2014 and 2019 [33].

While the literature identified multiple and often overlapping COVID-19-related stressors that adversely affected the mental health of parents and other caregivers of children, economic insecurity stands out as one of the strongest and most consistent sources of parent and caregiver stress [12,24,25,28,42,46–49]. For example, using data from 2,174 Bangladeshi mothers with children aged 0–5 years old, Hamadani et al. [25] found that 96% of respondents experienced a reduction in paid work during a COVID-19 lockdown, with average monthly family income falling from US$212 to US$59, and food insecurity increasing by more than 50%. Self-reported maternal depressive symptoms, as measured by the Centre for Epidemiologic Studies-Depression Scale (CESD-D), significantly increased during the lockdown period compared to before the pandemic; and nearly 70% of the sample reported that their anxiety had risen during the lockdown. A US study of 561 parents and other caregivers of young children (aged 2–7 years old) and who were hourly workers found that the majority of respondents reported a loss of household income (69%) and even job loss (60%), which linked with increased anxiety, depression, anger, and irritability, as well as worse sleep quality [24].

Numerous studies found that parent and caregiver stress, depression, and anxiety during the COVID-19 pandemic was associated with reduced responsive caregiving, including, for example, lower parent–child closeness and more avoidant or harsher parenting attitudes and behaviours [10,13,16,27,28,32,37,38,41,42,48]. A New Zealand study found that parents of young children (aged 4–5 years old) who experienced more significant depressive symptoms and perceived stress during a COVID-19 lockdown reported harsher parenting and less warm/responsive parenting, as measured by the Parenting Styles and Dimensions Questionnaire (PSDQ); and parent–child relationships were also of lower quality [32]. A US study of low-income families with preschool-age children found that parental job loss and income loss during the pandemic were strongly associated with parents’ depressive symptoms, stress, diminished sense of hope, and negative parent–child interactions, including losing their temper or yelling at their child [27]. However, parents who lost their job but did not experience corresponding household income decline, perhaps due to economic stimulus relief for low-income families, were more likely to report enjoying spending time with their child during the COVID-19 pandemic and engaging in positive parent-child interactions, such as playing with or hugging their child, compared to those who experienced a decline in household income. These results suggest that efforts to ensure economic security may help address parents’ and other caregivers’ mental health needs and reduce adverse parenting risk. Studies also noted more couple conflict [50] and increased substance use [51] among parents and caregivers experiencing stress and anxiety during the pandemic.

Few studies to date have discussed potential interventions to support parents and other caregivers with COVID-19-related stress, anxiety, and mental health issues or ways to protect parents’ capacity for responsive caregiving during the crisis. One study found mothers rarely utilized mental health services during the COVID-19 pandemic due to various barriers, such as a lack of time or energy, cost of service, not believing they needed such services, and uncertainty on access [14]. Studies found that partner support and cooperative co-parenting [13,14,20,32] and moderate physical exercise [17, 29] were associated with better mental health and coping among parents and caregivers during the pandemic. Thus pandemic-related strategies for parents and caregivers could focus on supportive parenting interventions and home or internet-based exercise interventions. Lastly, a few studies point towards mixed and potentially positive impacts of the COVID-19 restrictions on deepening family ties, improving couples’ relationships, and developing new family hobbies or activities [13,22,46,52–54]. However, such positive benefits were typically found among the parents and caregivers of older children or adolescents and among families with several protective factors, including financial stability, comfortable and safe homes, enough food to eat, good health, and healthy couple relationships [22, 46].

### Breastfeeding support during COVID-19

Breastfeeding not only provides an infant with nutrition, but it is also a process of responsive caregiving in which the mother and her young child interact closely in a safe and secure relationship. Five studies related to breastfeeding support during the pandemic indicate that lockdowns resulted in reduced breastfeeding support and care [55–59]. For example, over a quarter of respondents in a national U.K. study struggled to access breastfeeding support during the pandemic, and some stopped breastfeeding before they intended to [55]. Among the women who stopped breastfeeding during the pandemic, the most common reason was the lack of breastfeeding support and face-to-face help with practical problems, such as latching. Some valued the virtual health appointments among the women who had to shift from face-to-face lactation support to digital consultations. In contrast, others reported feeling violated during this impersonal interaction, describing how they felt discomfort recording their infant trying to latch onto the breast [59]. During the pandemic, some breastfeeding women reported some positive benefits, including having more time to focus on breastfeeding and responsive feeding, with fewer visitors, more, privacy and more significant breastfeeding support from fathers [58].

### Fathers’ engagement in childcare and gendered employment patterns

The evidence suggests that increases in fathers’ time at home during COVID-19 have reduced gender gaps in childcare division, which may have promoted positive changes to family dynamics and more similar caregiving arrangements. In total, eight studies reported that fathers took on more childcare responsibilities during the COVID-19 pandemic [60–67]. For example, a survey of 1,536 Australian parents of children (aged 0–16 years old) reported that fathers averaged 2.21 daily hours on active childcare before the pandemic, and this average increased to hours during the pandemic, thereby narrowing the childcare gap between mothers and fathers [62]. However, mothers continued to carry the burden of extra childcare and household responsibilities during the pandemic [68], and gendered employment patterns were exacerbated [60,67,69–71]. In particular, mothers were more likely than fathers to lose their jobs during the crisis, spend less time on paid work, and be interrupted during work hours, principally by childcare [1]. There is an absence of studies examining how parents and other caregivers of young children, including infants and toddlers, negotiate childcare responsibilities and household work during crises. There was also a lack of studies discussing male caregivers’ perspectives and experiences of caregiving during the pandemic and quality of interaction and exploring caregiving responsibilities and arrangements of single parents.

### Opportunities for early learning during COVID-19

Evidence on home-based early learning during the COVID-19 pandemic is limited compared to the other thematic areas in the review. What is available indicates that many children in early childhood education did not have contact with teachers or access to remote early learning during school closure [66,72,73]. For example, a study of Ethiopian parents and caregivers of young children found that only 10% of children who enrolled in pre-primary had been in contact with teachers or school principals, with significant differences found by household wealth and across regions [73].In this study, half of parents and caregivers reported supporting educational or learning activities for pre-primary children, including 12% who had used radio lessons while schools were closed. Children living in rural or remote areas had significant disadvantages regarding access to electricity, technologies, and children’s books or learning materials.

Other studies provide evidence of frustration and negative attitudes among caregivers about distance learning benefits and values, especially for preschool-aged children [74–76]. Home-based learning has been challenging for young children for various reasons, including limited learning materials and technological barriers [77, 78]. Children who were looked after by grandparents faced challenges due to their limited awareness about online technologies [77]. Parents noted concerns about lack of social interaction, sedentary behaviour, and limited physical activity during school closures [21,79,80]. A study of parents of children (aged 2-4 years) reported that almost half of the children spent no time playing with another child in their household during lockdowns [21]. Evidence is limited and mixed on the nature and extent of caregiver engagement in adult-child reading, playful parenting, and other early learning activities during the lockdown. Some studies find that parents engaged more in such activities during the lockdown [66, 73] while others reported they engaged less than before the pandemic [72]. Parents who met the criteria for probable major or severe depression and parenting stress were negatively associated with parents’ perceived preparation to educate at home [79].

### Children’s play and physical activity during COVID-19

Nine studies reported that COVID-19 restrictions, such as the closure of schools and playgrounds, the cancellation of sports and activity classes, and reduced social interactions with peers, were linked to reduced outdoor play and physical activity and more sedentary behaviour among children [72,81–88]. One study found that nearly 9 out of 10 Canadian parents reported increased screen time since the beginning of the pandemic among young children aged 18 months old to 5 years old [81]. More than half of the parents in this study reported that children’s physical activity decreased during the pandemic, citing challenges due to the lack of indoor space and the variety of available toys. Another study found a dramatic decline in children’s outdoor physical activity and much higher screen time use (average of 5 hrs/day) during the initial period of the COVID-19 outbreak, compared with before the restrictions [83]. Play indoors did not seem to replace active play outdoors, resulting in a net decline in children’s play-based activities. Cities and areas with the highest number of COVID-19 cases, and thus most stringent restrictions, had the most significant drop in children’s outdoor play [84]. Living in houses (as opposed to apartments) was correlated with increased outdoor activities [86].

### Child protection from violence and neglect during COVID-19

In terms of children’s safety and security, evidence indicates that COVID-19 has reduced child maltreatment referrals. Thirteen studies across seven countries, mostly utilizing publicly available administrative data, suggest that referrals of potential abuse, neglect, and maltreatment to child protective services were substantially lower during the COVID-19 pandemic than before quarantine measures were put in place [89–101]. Surveys of child protection workers similarly reported decreased referrals and more difficulty identifying children and families in need during lockdown [99,102,103]. For example, a study based on data from January 2015 to May 2020 for New York City’s Administration for Children’s Services reports there were 29% fewer allegations of child maltreatment for March 2020—when school closures first began— compared to previous years [96]. Substantially fewer allegations of child maltreatment were reported by education personnel and other mandated reporters, including childcare personnel and medical health personnel during this time. Non-mandated reporters, including family members and neighbours, also made fewer referrals, which may be linked to social distancing and limited contact between children and those outside their households. Bullinger et al. (2020a, 2020b) found that in Chicago and Georgia in the United States, emergency calls for domestic crimes rose in March and April 2020 but calls for potential child abuse fell, potentially due to the lessened visibility of child victims than adults.

These changes do not reflect an actual reduction in the incidence or prevalence of maltreatment. The decrease in referrals is likely due to the given lockdown measures that preclude children from having regular contact with educators and other mandated reporters. Although many school districts have transitioned to online learning during the COVID-19 pandemic, evidence suggests that educators typically receive inadequate education related to recognizing and responding to child maltreatment. Virtual formats for teaching do not lend themselves readily to the identification of child maltreatment. With reduced exposure to schools and other social services, there is widespread concern that child maltreatment victims are going undetected.

Two studies indicate a striking rise in child abuse-related injuries among babies and toddlers during the pandemic [104, 105]. Comparing cases of children’s head trauma caused by suspected abuse, there were 10 cases at an urban medical facility in the United Kingdom over the March–April 2020 period, representing an increase of 1493% compared to the previous three years preceding the lockdown [105]. Although alarming, it is important to consider that COVID-19 has led to significant healthcare delivery changes, and many centres have diverted non-COVID-related health emergencies to specific healthcare facilities, which may account for the increases in abuse head trauma at particular institutions. One additional study examined the association between COVID-19 and potential child abuse and found that COVID-19 job loss among parents and caregivers was significantly associated with psychological maltreatment, including verbal threats, belittling, and ridiculing, with larger effects for younger children [106]. Given previous evidence detailing the strong links between caregiver stress and the risk of child maltreatment, the economic-related impacts of COVID-19 may contribute to elevated rates of child maltreatment; however, this relationship has not yet been substantiated.

### Child injuries during COVID-19

Data on emergency department visits and injuries suggest that children’s environments have shifted during the pandemic and present different child safety threats. Nine studies from eight countries reported a significant reduction in emergency department presentations and operations for injuries among children during government-imposed lockdown restrictions, especially related to admissions and operations for sporting-related injuries and those that occur on playgrounds bans on sports and access to playgrounds [107–115]. For example, a study based on data from a large acute paediatric hospital in London, UK, found that the prevalence of referrals for child injuries was reduced by nearly two-thirds during the lockdown from 17 March to 28 April 2020, compared to in the same period in 2019 [111]. In contrast, some centres found a relative increase in injuries, including bicycle injuries [107, 109], burn injuries [116], ingestions and poisonings [108], and electrical injuries [114]. The higher incidence of child burn injuries during COVID-19 lockdown was equivalent to the percentage of admissions during a typical summer break and suggests that child supervision may be a significant factor during school closures [116]. The general demographic of those presenting with injuries also changed during the pandemic compared to before the pandemic, with a significantly younger median age [111,113,115]. Parents and other caregivers may have treated more minor injuries at home or, in some cases, may have avoided seeking medical attention due to the concern of contracting COVID-19 in hospital facilities. Physical distancing measures, including a ban on sports and children’s use of playgrounds during lockdowns, is likely linked to the overall reduction in child injuries seen through emergency room admissions. At the same time, there may have also been a shift in care-seeking behaviour, with parents and caregivers being more anxious about attending hospital due to the risk to themselves and the child of contracting COVID-19.

## DISCUSSION AND CONCLUSION

Emerging evidence from the COVID-19 pandemic, mainly drawn from parents and other caregivers’ surveys, shows striking similarities in how essential components of nurturing care –responsive caregiving, support for young children’s learning, and children’s safety and security-have been disrupted. While there are some reports of unexpected positive benefits of the pandemic on families, such as increased involvement in caregiving among fathers, more commonly, the studies’ findings reveal issues of concern, with parent and caregiver mental health difficulties and strained parent–child relationships, increased screen time among children, limited opportunities for outdoor play, and fractured systems for responding to potential child neglect and maltreatment. As with many other features of this pandemic, not all families are affected equally. Those with salaried jobs are far less likely to experience income loss than those with informal, daily wage jobs. Social safety nets such as stimulus packages for families may reduce parenting stress brought about by COVID-19-related economic hardship and reduce the risk of harsh and non-responsive parenting. The pandemic is disproportionately affecting parents and other caregivers with pre-existing mental health conditions and caregivers of children with disabilities. Families with young children, particularly mothers, may be disproportionately affected by childcare and school closures, the reduced ability to undertake paid work, and increased caregiving demands.

Based on these findings, we suggest policy and programme responses for action by governments, civil society, international and community-based organizations (see Box 1). All of these options should be pursued vigorously and simultaneously so that the pandemic does not break the continuity nor lower the quality of provision of nurturing care and wipe out decades of progress, especially for low-income families and children. These approaches must be coordinated across sectors and aligned with practical social safety support to families and other caregivers of young children who experience socioeconomic hardship.

### Box 1: Key programme and policy implications

1. **Scale-up of social protection mechanisms to ensure families can meet their basic income needs and food and housing security.** Governments should reflect on, and rebalance, expenditures going to families and children. Supporting parent and caregiver mental health and minimizing family stress should continue to be a top priority in COVID-19 response plans and prevention-based approaches. There remains a huge unmet need to expand and diversify social protection responses, including the immediate delivery of food packages, longer-term paid parental leave, accessible childcare, universal basic income, and waivers for utility bills and rent or mortgage payments, to avoid further indebtedness or evictions. Families are much less likely to benefit from family and parenting support when their basic needs are not met.
2. **Improve access to and use of mental health and psychosocial support services for parents and other caregivers of young children are essential**. Digital interventions for anxiety and depression might include information provision, connectivity and triage, automated and blended therapeutic interventions (such as apps and online programmes), and telephone calls and home visits, especially for those without connectivity. Given the need to disseminate affordable mental health support widely due to COVID-19, group-based telehealth interventions may allow for an increased reach of psychological services in a time of elevated need.
3. **More evidence is needed on scalable responsive caregiving interventions, responsive caregiving, early learning activities and play, appropriate for the conditions of physical distancing and lockdowns.** Programs that foster peer support and online platforms should be evaluated to ensure acceptable efficacy in supporting caregivers with young children, particularly on aspects of early learning and play. Guidance and resources to help parents with balancing screen time with physically active, screen-free activities may be warranted.
4. **Child protection systems should be redesigned**, and educators trained to identify signs of child abuse and neglect specific to a distance-learning model. Home visitation programmes, with appropriate protection of social service staff, should continue to the extent possible, particularly with high-risk families. To increase awareness and access to reporting hotlines and other maltreatment reporting resources, policy makers may consider a variety of dissemination means, including public signage, as well as broadcast and social media.

A recent report of case studies of programme implementation to support nurturing care during COVID-19 illustrates several innovative examples of how organizations in various contexts have adapted to support nurturing care efforts during COVID-19 [117]. For instance, programmes have organized emergency food delivery services for vulnerable families. Others have developed new training materials for frontline workers that integrate both mental health and nurturing care components. Organizations concerned with children’s health and development in many countries have implemented virtual caregiver support meetings, telephone or internet-guided learning activities for young children, and online stress reduction resources for parents and other caregivers.

The evidence in this rapid review was primarily collected during the pandemic’s first stage of lockdowns, representing only the early phase of the pandemic. Future research must investigate the longer-term impact of the COVID-19 pandemic on families as we enter a period of a gradual relaxation of lockdown measures in some regions and second waves in others. Gaps in the evidence base remain, with a lack of studies in low- and middle-income countries. There is an urgent need for action-oriented implementation studies – those that go beyond identifying trends and begin to pinpoint “what works” to effectively promote and protect nurturing care during health emergencies such as COVID-19. In particular, there is a need to identify effective interventions and strategies for families experiencing income loss, food insecurity, mothers with young children, families with disabled children, and those with pre-existing mental health challenges. Also, further inquiry is needed into the effects of COVID-19 lockdowns on early learning and children’s play, which is transformative for children, allowing them to creatively develop their imagination, dexterity, and growth. The relative absence of studies related to early learning and playful parenting indicates a need for additional evidence on the nature and prevalence of early learning and playful parenting and innovative ways to protect and promote them during health emergencies.

## Data Availability

The data supporting the findings of this study are available within the article and its supplementary materials.

## SUPPLEMENTARY TABLE: SUMMARY OF INCLUDED STUDIES

**Responsive caregiving: Caregiver stress and mental health**

Responsive caregiving: Caregiver stress and mental health
| Reference | Date Released | Setting | Population | Study Design | Data Source | Summary of Findings |
| --- | --- | --- | --- | --- | --- | --- |
| **Abdelrahman, M. K., et al.** Impact of social distancing on the mental health of parents and children in Qatar. | Sept-20 | Qatar | *N* = 308<br>Parents of children <18 years | Cross-sectional | This online survey, conducted between June 11 and July 10, 2020, explores parents' mental health and coping strategies during COVID-19. | Results showed a significant positive correlation between social distancing practices and parents' activities with children and their coping strategies, suggesting that staying at home for an extended period and not going outside unless necessary motivate parents to develop activities for the children, |
| **Adibelli, D., and A. Sumen.** The Effect of the Coronavirus (Covid-19) Pandemic on Health-Related Quality of Life in Children. | Oct-20 | Turkey | *N* = 597<br>Parents of children ages 7-13 years | Cross-sectional | The online survey, conducted between March 30 and April 20, 2020, explored the relationship between parents' mental health and children's quality of life during the pandemic. | 45% of parent respondents reported that staying at home due to the pandemic negatively affected their mental health, and 17% reported a positive affect on their mental health. Quality of life scores (i.e., emotional well-being, family, friends) were lower among children of the parents who were fear/anxiety about coronavirus. |
| **ARNEC.** A survey on Perspectives on the impact of COVID-19 on young children and early childhood development in the Asia-Pacific region. | Jun-20 | Multiple<br>N=30 | *N* = 684<br>ECD Stakeholders | Cross-sectional | The survey, conducted between 17 April 2020 and 16 May 2020, explored perspectives of the impact of COVID-19 on families and young children in Asia Pacific. | More than 50% of ECD respondents reported that families are under pandemic induced stress and facing challenges in coping under the heightened stress. Income losses, school/ECD centre closures and prolonged home stay are top reasons for family stress. |
| **Asbury, K., et al.**<br>How is COVID-19 Affecting the Mental Health of Children with Special Educational Needs and Disabilities and Their Families? | Jul-20 | UK | *N* = 241<br>Parents of children ages 5-18 with Special Educational Needs and Disabilities | Qualitative | In an online survey conducted between March 22-April 1, 2020, participants were asked to answer the following free response question: Please describe in your own words how the coronavirus outbreak is affecting your mental health and your child's mental health. | Both parents and children appear to be experiencing loss, worry and changes in mood and behaviour as a result of the rapid social changes that have occurred. Some parents reported feeling overwhelmed and described the impact of child understanding and awareness. Finally, a minority of parents reported that COVID-19 has had little impact on mental health in their family, or has even led to improvements. |
| **Brown, S. M., et al.**<br>Stress and parenting during the global COVID-19 pandemic. | Aug-20 | USA | *N* = 183<br>Parents of children <18 years | Cross-sectional | An online survey, conducted from April 21 to May 9, 2020, to examine the relationships among COVID-19 risk factors, mental health risk factors, protective factors, parental perceived stress, and child abuse potential. | Greater COVID-19 related stressors and high anxiety and depressive symptoms are associated with higher parental perceived stress. Receipt of financial assistance and high anxiety and depressive symptoms are associated with higher child abuse potential. Conversely, greater parental support and perceived control during the pandemic are associated with lower perceived stress and child abuse potential. Results indicate racial and ethnic differences in COVID-19 related stressors. 10% reported a positive change: having more time with family. |
| **Cameron, E. E., et al.**<br>Maternal Psychological Distress & Mental Health Service Use during the COVID-19 Pandemic. | Jul-20 | Primarily Canada | *N* = 641<br>Mothers of children ≤8 years, including expectant mothers | Cross-sectional | An online survey, conducted between April 14 and April 28, 2020, described rates of maternal depressive and anxiety symptoms during the COVID-19 pandemic, and identified risk and protective factors. | In the full sample, 41.51% of mothers met the clinical cut-off scores of $\geq 13$ for EPDS and $\geq 16$ for CESD indicating depression. Over one-third of mothers (38.62%) indicated current financial strain. |
| **Carroll, N., et al.** The Impact of COVID-19 on Health Behavior, Stress, Financial and Food Security among Middle to High Income Canadian Families with Young Children. | Aug-20 | Canada | *N* = 254<br>Parents of children ages 18 months to 5 years | Cross-sectional | An online survey administered between 20 April 2020 to 15 May 2020 to explore impact of COVID-19 on physical activity, screen time, and sleep among parents and children, as well as stress, financial and food insecurity of families. | Parents reported moderately high levels of stress. Several parents reported coping strategies they use to manage stress and life changes due to COVID-19 restrictions including being more mindful and focusing on the positive aspects of having more time together as a family, taking prescription medication or seeking therapy, and following their pre-COVID-19 routines |
| **Ceulemans, M., et al.** Mental Health Status of Pregnant and Breastfeeding Women during the COVID-19 Pandemic: A Call for Action. | Jul-20 | Belgium | *N* = 5866<br>Pregnant and breastfeeding mothers | Cross-sectional | Online survey, limited details on sampling and recruitment, explored maternal mental health status using the Edinburgh Depression Scale and the Generalized Anxiety 7-item scale | The prevalence of self-reported major depressive symptoms (EDS $\geq 13$) in pregnancy (25.3%) and postpartum (23.6%) was higher compared to estimates obtained prior to the pandemic. Generalized anxiety during the lockdown, with more than 40% of the women surveyed scoring $\geq 5$ on the EDS subscale of anxiety. In addition, 14% met the criteria for high anxiety on the GAD-7 ($\geq 10$). |
| **Chu, K., et al.** Parenting Under Pressure | 2020<br>*Preprint* | USA | *N* = 56<br>Parents of children ages 6-17 years | Mixed methods | Thematic analysis to identify themes in parents' and children's reflective writing about COVID-19, conducted between April to May 2020. | The most common themes identified from parent reports surrounded concerns about the impact of COVID-19 on their children, health concerns for others, and the stressful balancing act of parenting, schooling, and working from home. In addition, many parents reported gratitude for what they had, and reflected on the upsides of the pandemic for family relations. |
| **Chung, G., et al.**<br>Associations Between Work-Family Balance, Parenting Stress, and Marital Conflicts During COVID-19 Pandemic in Singapore. | Jun-20<br>*Preprint* | Singapore | *N* = 258<br>Parents of children <12 years | Cross-sectional | An online survey disseminated from April 22, 2020, to May 5, 2020 explored work, family life and parenting during the pandemic. | Results indicated three profiles of Work-Family Balance (WFB) and social support: Profile 1 (Poor, *n* = 38, 19%); Profile 2 (Moderate, *n* = 76, 38%); and Profile 3 (Strong, *n* = 87, 43%). Parents' age, race, education, family income, any domestic helpers, and number of caregivers in the household did not predict membership in any of the profiles. Mothers were more likely than fathers to be in the Moderate and Poor profiles. Parents with older children were less likely to be in Profile 1 and 2. Parents who reported a greater impact of COVID-19 and/or with more children in the household were more likely to be in Profile 2 than in Profile 3. |
| **Davenport, M. H., et al.** Moms Are Not OK: COVID-19 and Maternal Mental Health. | Jun-20 | Mostly USA and Canada | *N* = 900<br>Expectant and mothers of children ≤12 months | Cross-sectional | An online survey, conducted between April 14 and May 8, 2020, explored the COVID-19 pandemic on the mental health and physical activity of pregnant and postpartum women. | An EPDS score >13 was self-identified in 40.7% of participants. Moderate to high anxiety (STAI-state score >40) was identified in 29% of women before the pandemic vs. 72% of women currently. |
| **Dhiman, S., et al.**<br>Impact of COVID-19 outbreak on mental health and perceived strain among caregivers tending children with special needs. | Oct-20 | India | *N* = 264<br>caregivers of children with special needs | Cross-sectional | Caregivers completed an online survey from April 29, 2020 to May 22, 2020 that assessed demographics, use and perspective on tele-rehabilitation, homecare therapy, caregiver's strain and mental health | Prevalence of depression, anxiety and stress symptoms were 62.5%, 20.5% and 36.4%, respectively. A significant difference in caregiver strain was observed during the outbreak and pre-outbreak, as measured retrospectively. Caregivers not using tele-rehabilitation along with a perception of it being a poor medium for rehabilitation were at greater risks for poor mental health, while negative perception on homecare therapy were strongly associated with higher psychological symptoms and strain. Working from home was a risk factor for poor mental health. Caregivers of children older than 6 were less prone for depression, anxiety and stress symptoms. |
| **Di Giorgio, E., et al.**<br>The Interplay between Mothers' and Children Behavioral and Psychological Factors during COVID-19: An Italian Study. | Aug-20 | Italy | *N* = 245<br>Mothers of children ages 2 to 5 years | Cross-sectional | An online survey, conducted between April 1-9, 2020 to explore both mothers' habits, behaviors, and emotions and those of their children during the quarantine, and retrospectively, before the national lockdown (i.e., in late February) | Most respondents (66.5%) had to stop working or to start working from home. A general worsening of sleep quality and distortion of time experience in both mothers and children, as well as increasing emotional symptoms and self-regulation difficulties in children, was observed. Moreover, even when the interplay between the behavioral and psychological factors was investigated, the factor that seems to mostly impact both mothers' and children's psychological well-being was their sleep quality. |
| **Dib, S., et al.** Maternal Mental Health and Coping during the COVID-19 Lockdown in the UK: Data from the COVID-19 New Mum Study. | Sep-20 | United Kingdom | *N* = 1329<br>Mothers of children $\leq 12$ months | Cross-sectional | An online survey May 27, 2020, and will remain open until at least December 31, 2020, explores predictors of maternal mental health and coping during COVID-19. | Support with her own health (95% confidence interval [CI] 0.004–0.235), contacting infant support groups (95% CI –0.003 to 0.252), and higher gestational age of the infant (95% CI 0.000–0.063) predicted better mental health. Support with her own health and more equal division of household chores were associated with better coping. |
| **Drouin, M., et al.**<br>How Parents and Their Children Used Social Media and Technology at the Beginning of the COVID-19 Pandemic and Associations with Anxiety. | Jul-20 | USA | *N* = 260<br>Parents of children $\leq 18$ years | Cross-sectional | Online survey data, collected from March 20-25, 2020, explored children's use of social media and technology during the COVID-19 pandemic and its association with anxiety. | 92% of parents and 82% of children had increased their technology use to connect with others. Bivariate correlations revealed significant positive associations between parent anxiety and parent technology use, social media use, and checking messages/ news, but not connecting via video or taking time for self. Those with greater financial preparedness and older children were able to take time for themselves more frequently, and those with older children connected with others via video less frequently than those with younger children. |
| **Evans, S., et al.** From “It Has Stopped Our Lives” to “Spending More Time Together Has Strengthened Bonds”: The Varied Experiences of Australian Families During COVID-19. | Oct-20 | Australia | *N*=2130<br>Parents of children 0-18 years | Qualitative | During an online survey conducted between April 8 and April 28, 2020, participants were asked to respond via an open-ended question about how COVID-19 had impacted their family. | Six themes were derived from the data, including “Boredom, depression and suicide: A spectrum of emotion,” “Families are missing the things that keep them healthy,” “Changing family relationships: The push pull of intimacy,” and others. Messages around loss and challenge were predominant, with many reporting mental health difficulties and strained family relationships. However, not all families were negatively impacted by the restrictions, with some reporting positive aspects, including opportunities for strengthening relationships, finding new hobbies, and developing appreciation, gratitude, and tolerance. |
| **Farewell, C. V., et al.** A Mixed-Methods Pilot Study of Perinatal Risk and Resilience During COVID-19. | Jul-20 | USA | *N* = 31<br>Mothers of children ≤6 months and expectant mothers | Mixed methods | Mixed methods study conducted between March and April 2020 explored mental health and well-being during the perinatal period. | New moms shared that social isolation was significantly affecting their postpartum mental health. Positive impacts related to the COVID-19 pandemic, included being able to work from home during pregnancy, Increased connection and bonding with their immediate family unit, partner supports in the home to share caregiving responsibilities; and increased access to remote postnatal and postpartum care. |
| **Gassman-Pines, A., et al.** COVID-19 and Parent-Child Psychological Well-being. | Oct-20 | USA | *N* = 561<br>Parents/ caregivers of children ages 2 to 7 years | Longitudinal | Daily survey data were collected between February 20 and April 27, 2020, from hourly service workers with a young child in a large US city. A subsample of 561 completed a one-time survey about the effects of the crisis between March 23 and April 26. | Ordered probit models revealed that the frequency of parent-reported daily negative mood increased significantly since the start of the crisis. Many families have experienced hardships during the crisis, including job loss and income loss. Both parents’ and children’s well-being was strongly associated with the number of crisis-related hardships. |
| **Günther-Bel, C., et al.** A Mixed-method Study of Individual, Couple, and Parental Functioning During the State-regulated COVID-19 Lockdown in Spain. | Jul-20 | Spain | *N* = 407 Adults including parents | Mixed methods | An online survey conducted between March 24 and April 7, 2020 explored individual and family well-being during the first 3 weeks of state-regulated lockdown. | An online survey conducted between March 24 and April 7, 2020, described perceived improvement as family connection, better communication, emotional expressiveness, teamwork spirit, and balance between individual and shared needs. The overall prevalence of improvement themes exceeded the prevalence of deterioration themes, mainly conflict atmosphere and unbalanced needs (most likely with children with children at home). Deterioration themes were associated with the presence of preschoolers. |
| **Hamadani, J. D., et al.** Immediate Impact of Stay-at-Home Orders to Control COVID-19 Transmission on Socioeconomic Conditions, Food Insecurity, Mental Health, and Intimate Partner Violence in Bangladeshi Women and Their Families: An Interrupted Time Series. | Aug-20 | Bangladesh | *N* = 2417 Mothers of children ≤5 years | Quasi-experimental | An interrupted time series was used to compare data collected on income, food security, and mental health a median of 1 year and 2 years before the COVID-19 pandemic to data collected during the lockdown. | Mothers' depression and anxiety symptoms increased during the lockdown. Among women experiencing emotional or moderate physical violence, over half reported it had increased since the lockdown. |
| **Hiraoka, D., and A. Tomoda.** Relationship between Parenting Stress and School | Jun-20 | Japan | *N* = 353 Parents of children <18 years | Cross-sectional | An online survey, conducted between 29 and 30 April 2020, administered the Parenting Stress Index – | Parental stress was significantly higher after school closures, compared to parental stress before school closures, based on recall ($t = 4.89$, $p < .01$, $d = 0.12$). |
| Closures Due to the COVID-19 Pandemic. |  |  |  |  | Short Form (PSI-SF) to measure parenting stress during the pandemic. |  |
| **Joyce, Kayla M., et al.** Changes in Maternal Substance Use During the COVID-19 Pandemic. | 2020<br>*Preprint* | Canada | *N* = 508<br>Mothers of children ≤8 years | Cross-sectional | Online survey data collected April 14th to 28th 2020 to identify associations between mental health and substance use. The EPDS and GAD-7 were completed. | Of participating mothers, 54.9% did not change their substance use, 39.2% increased their use, and 5.9% decreased their use. Findings indicate that mothers with an anxiety disorder or clinically relevant anxiety symptomatology were more likely to report increased substance use during the COVID-19 pandemic. |
| **Kalil, A., et al.** Impact of the COVID-19 Crisis on Family Dynamics in Economically Vulnerable Households. | Oct-20 | USA | *N* = 572<br>Parents of preschool-age child | Cross-sectional | A telephone survey, conducted from May 3 to July 20, 2020, to explore parental mental health and parent-child interaction during the pandemic. | Parental job and income losses are strongly associated with parents' depressive symptoms, stress, diminished sense of hope, and negative interactions with children. Parents who experience a job and income loss, as well as those with just income loss, are more likely to lose their temper or yell at their child. Parents who have job and income loss are more likely to be regular readers with their children. |
| **Lee, S. J., et al.** Parenting Activities and the Transition to Home-Based Education During the COVID-19 Pandemic. | Oct-20<br>*Preprint* | USA | *N* = 405<br>Parents with children <12 years | Mixed methods | An online survey, including open-ended questions, conducted from April 24-25, 2020, examined parental mental health and the transition to home-based learning. | 78% of parents said they were educating their child at home due to COVID-19. Two out of every five parents met the PHQ-8 criteria for major depression or severe major depression (40.0%) and the GAD-7 criteria for moderate or severe anxiety (39.9%). Multivariate analyses indicated that, compared to non-depressed parents, parents who met criteria for probable major or severe depression and parenting stress were negatively associated with parents' perceived preparation to educate at home. Compared to parents with minimal or mild anxiety, parents with moderate or severe anxiety reported higher child anxiety. Content analyses of open-ended questions indicated that school closures were a significant disruption, followed by lack of physical activity, and social isolation. |
| **Limbers, C. A., et al.**<br>Physical Activity Moderates the Association between Parenting Stress and Quality of Life in Working Mothers during the COVID-19 pandemic. | Oct-20 | USA | *N* = 200<br>Full-time working mothers of children ≤5 years | Cross-sectional | Online survey of mothers completed during April 6th and April 13th, 2020, to examine physical activity, parenting stress and quality of life during the pandemic. | Conducted from results showed that greater parenting stress was associated with poorer maternal quality of life (physical health, psychological, social relationships, and environment quality of life), after controlling for socio-demographic variables. Multiple linear regression analysis suggest that moderate intensity physical activity may be one mechanism that attenuates the association between parenting stress and diminished social relationships quality of life during. |
| **Marchetti, D. et al.**<br>Parenting-Related Exhaustion During the Italian COVID-19 Lockdown. | Oct-20 | Italy | *N* = 1,226<br>Parents of children ≤13 years | Cross-sectional study | An online survey administered between April 3 and 14 2020 to explore emotional exhaustion, parental resilience, social connections, and psychological distress during the lockdown. | Results indicate that more than 80% of parents reported high levels of psychological distress, and nearly one in five (17%) reported significant parenting-related exhaustion, with mothers more severely affected. Multiple regression analyses showed that greater parenting-related exhaustion was predicted by psychological distress, lower parental resilience, fewer perceived social connections, and being single, as well as having a child with special needs, having a large number of children, and having younger children. |
| **Mazza, C., et al.** How Personality Relates to Distress in Parents during the Covid-19 Lockdown: The Mediating Role of Child's Emotional and Behavioral Difficulties and the Moderating | Aug-20 | Italy | *N* = 833<br>Parents of children ages 3–13 years | Cross-sectional | An online survey was administered to 833 participants from 3 to 15 April 2020 to investigate psychological distress of parents during the lockdown and contributing factors. | For parents living only with child(ren), high levels of psychological distress depended exclusively on their levels of neuroticism. For parents living with at least one other person in addition to child(ren), distress levels were also mediated by child behavioral and emotional difficulties. Motherhood emerged as a significant factor contributing to greater distress. Furthermore, parent psychological distress decreased in line with increased child age. |
| Effect of Living with Other People. |  |  |  |  |  |  |
| **McRae, C. S., et al.**<br>Parents' Distress and Poor Parenting during COVID-19: The Buffering Effects of Partner Support and Cooperative Coparenting. | 2020<br>*Preprint* | New Zealand | *N* = 362<br>Parents of children ages 4 to 5 years | Longitudinal | An online survey conducted from March 26 to April 28, 2020 assessed harsh parenting, warm/responsive parenting, and parent-child relationship quality among a cohort of parents who had already been surveyed pre-pandemic. | Parents who experienced greater distress during the lockdown reported harsher parenting, less warm/responsive parenting and lower parent-child relationship quality. Greater psychological distress predicted residual increases in harsh parenting when parents perceived low partner support, but the detrimental effects of parents' distress was buffered when parents perceived high levels of partner support. |
| **Nolvi, S., et al.**<br>Parental Depressive and Anxiety Symptoms as a Response to the COVID-19 Pandemic: A Birth Cohort Follow-up Study. | 2020<br>*Preprint* | Finland | *N* = 651<br>Parents of children ages 5-8 years | Longitudinal | Based on a sample from a longitudinal cohort study, the FinnBrain Birth Cohort Study, data collected from May 4 and June 7, 2020 using the EPDS, Symptom Checklist 90 | Higher symptom level was associated with COVID-related stressors, being mother (vs. father), prior symptoms and prior negative life events. Importantly, there was a significant increase in both depressive and anxiety symptoms and in subjects scoring above the screening cut-off of depression from pre-pandemic to pandemic. The increase in symptoms more pronounced in mothers versus fathers. |
| **Oskovi-Kaplan, Z. A., et al.** The Effect of COVID-19 Pandemic and Social Restrictions on Depression Rates and Maternal Attachment in Immediate Postpartum | Sep-20 | Turkey | *N* = 223<br>Mothers, 48 hrs post-partum | Cross-sectional study | The survey was conducted in June 2020. The low-risk term pregnant women who gave birth were given the surveys Edinburgh Postpartum Depression Scale (EPDS) and Maternal Attachment | Conducted in June 2020, the median score obtained from the EPDS was 7 and 15% of the women were determined to have a risk for postpartum depression. The authors suggest the postpartum depression risk during the COVID-19 pandemic was increased when compared with previous reports in Turkey, while it was still lower than the studies reported from other countries. |
| Women: A Preliminary Study |  |  |  |  | Inventory (MAI) within 48 h after birth. |  |
| **Patrick SW, et al.**<br>Well-being of Parents and Children During the COVID-19 Pandemic: A National Survey. | Oct-20 | USA | *N* = 1011<br>Parents of children <18 years | Cross-sectional | As national online survey conducted from June 5-10, 2020 to determine how the pandemic and mitigation efforts affected the physical and emotional well-being of parents and children. | Since March 2020, 27% of parents reported worsening mental health for themselves, and 14% reported worsening behavioral health for their children. 48% reported loss of regular childcare, 16% reported change in insurance status, and 11% reported worsening food security. |
| **Peltz, J. S., et al.**<br>Mediators of the Association between COVID-19-Related Stressors and Parents' Psychological Flexibility and Inflexibility: The Roles of Perceived Sleep Quality and Energy. | Jul-20 | USA | *N* = 1003<br>Parents of children ages 5 to 18 years | Cross-sectional | Online survey conducted from March 27th to the end of April 2020 to explore COVID-19 related stressors, and | Path analyses suggested that higher levels of health-related stress were associated with lower levels of energy and lower perceived sleep quality, both of which were associated with higher levels of psychological inflexibility. In secondary analyses on the specific dimensions of flexibility and inflexibility, results suggested that lower energy levels indirectly linked health-related stress to lower levels of all 6 dimensions of flexibility and poorer sleep quality indirectly linked health-related stress to higher levels of self-as-content, fusion, and inaction. |
| **Romero, E., et al.**<br>Testing the Effects of COVID-19 Confinement in Spanish Children: The Role of Parents' Distress, Emotional Problems and Specific Parenting. | Sep-20 | Spain | *N* = 940<br>Parents of children ages 3 to 12 years | Cross-sectional | Online survey conducted in April 2020 to examine parenting distress and specific parenting practices (e.g., structured or avoidant parenting). | Data collected in April 2020 show that higher levels of parenting distress were significantly related with lower focused and structured parenting, and higher avoidant parenting. parenting distress was also affected by COVID-related stressors, including close contagion, keeping up with jobs and working at home. |
| **Roos, L. E., et al.**<br>Supporting Families to Protect Child Health: Parenting Quality and Household Needs During the COVID-19 Pandemic. | Sep-20 | Canada | *N* = 656<br>Parents or caregivers of children ages 1-8 years | Cross-sectional | Mixed methods were applied to data collected between April 14th - 28th, 2020 from the “Parenting During the Pandemic” survey. Data collection occurred through REDCap. | Caregiver depression, higher child parity, unmet childcare needs, and relationship distress predicted lower-quality parenting (i.e., more stress, fewer positive behaviors, and more negative behaviors). Caregiver depression was the most significant predictor across every parenting dimension, with analyses indicating medium effect sizes. Qualitative findings highlighted severe strains on parent capacities including managing psychological distress, limited social supports, and too much unstructured time. |
| **Russell, B. S., et al.**<br>Initial Challenges of Caregiving During COVID-19: Caregiver Burden, Mental Health, and the Parent–Child Relationship. | Aug-20 | USA | *N* = 420<br>Parents of children <18 years | Cross-sectional | Online survey conducted April 27–28, 2020s to explore family experiences and coping during the COVID-19 pandemic. | Caregiver burden was positively associated with generalized anxiety, depression. Male caregivers reported higher rates overall than female caregivers. Depression was significantly positively associated with child parent relationship conflict but not child parent relationship closeness. Caregiver burden was not significantly directly associated with child parent relationship conflict or child parent relationship closeness. Generalized anxiety did not significantly predict child parent relationship conflict or child parent relationship closeness. |
| **Spinelli, M., et al.**<br>Parenting Stress During the COVID-19 Outbreak: Socioeconomic and Environmental Risk Factors and Implications for Children Emotion Regulation. | Sep-20 | Italy | *N* = 810<br>Parents of children ages 2–14 years | Cross-sectional | Online survey from April 2nd to 7th, 2020 to examine level of household chaos, parenting stress, parent involvement during COVID-19. | Parents reporting higher levels of stress were less engaged with their children, they were less interested in children emotional well-being, they paid less attention to the child, and in general spent less time with the child. For SES at-risk families, the impact of parenting stress and involvement on children regulation strategies was stronger, with a protective role played by parental involvement on children’s negativity not evident for SES no-risk families. The majority of these families in our sample were characterized by a reduction of their income due to the loss of the job because of the pandemic. |
| **Spinelli, M., et al.**<br>Parents' Stress and Children's Psychological Problems in Families Facing the COVID-19 Outbreak in Italy. | Jul-20 | Italy | *N* = 854<br>Parents of children ages 2-14 years | Cross-sectional | Online survey conducted from April 2nd to 7th, 2020 to explore perceptions of individual and parent-child dyadic stress, and their children's emotional and behavioral problems. | Parents who reported more difficulties in dealing with quarantine show more stress. This, in turn, increases the children's problems. Living in a more at-risk area, the quality of the home environment, or the relation they have with the pandemic consequences, do not have an effect on families' well-being. |
| **Westrupp, E., et al.**<br>Child, Parent, and Family Mental Health and Functioning in Australia During COVID-19: Comparison to Pre-pandemic Data. | Sep-20<br>*Preprint* | Australia | *N* = 2365<br>mothers and fathers of children 0-18 years | Repeat cross-sectional | Online survey assessing mental health, substance use, couple conflict, parenting, and family functioning during 'stage three' COVID-19 restrictions in April 2020 was compared to pre-pandemic data from four population-based surveys. | Compared to pre-pandemic estimates, during the pandemic period parents reported higher rates of parent mental health symptoms, higher parenting irritability, lower family positive, and higher alcohol consumption. In multivariable analyses, pre-existing financial deprivation and COVID-19 stressors were associated with greater severity in parent and child mental health symptoms, parent emotion dysregulation, parenting irritability, couple conflict and family positive/ negative expressiveness. Parents and children with pre-existing mental health conditions had elevated difficulties during the pandemic across most domains. |
| **Willner, P., et al.**<br>Effect of the COVID-19 pandemic on the mental health of carers of people with intellectual disabilities. | Sep-20 | UK | *n* = 107<br>carers of adults with intellectual disability;<br>*n* = 100<br>carers of children <18 years with intellectual | Cross-sectional | An online survey conducted between 28 April and 5 June 2020 estimated whether the extent of mental health problems was greater than would have been seen under pre-pandemic | Relative to carers of children without intellectual disability, carers of both children and adults with intellectual disability had significantly greater levels of a wish fulfilment coping style, defeat/entrapment, anxiety, and depression. Differences were 2–3 times greater than reported in earlier pre-pandemic studies. Positive correlations were found between objective stress scores and all mental health outcomes. Despite |
| | | | disability; and<br>$n = 37$ carers of children <18 years without intellectual disability | | conditions, by comparing current data to earlier literature. The instrument assessed demographics, coping strategies, social support, anxiety, feelings of defeat/entrapment, and stress. | their greater mental health needs, carers of those with intellectual disability received less social support from a variety of sources. |
| **Xu, Y., et al.** Material Hardship and Parenting Stress among Grandparent Kinship Providers during the COVID-19 Pandemic: The Mediating Role of Grandparents' Mental Health. | Aug-20 | USA | $N = 362$ Grandparent caregivers of children < 18 years | Cross-sectional | An online survey conducted in June 2020 to examine material hardship, parenting stress, and grandparents' mental health. | A total of 28.18 % ($n = 102$) of grandparents agreed that they experienced parenting stress during the COVID-19 pandemic. A comparison between grandparent kinship providers that experienced parenting stress and those that did not found that the latter had a significantly lower score on experiencing material hardship. Similarly, grandparents that did not report parenting stress had significantly better mental health status. |
| **Yang, Y., et al.** How did COVID-19 Impact the Lives and Perceived Well-being of Parents? Using the Case of Singapore to Investigate the Mechanisms. | Oct-20<br>*Preprint* | Singapore | $N = 505$ adults caring for children <18 years from 198 families | Cross-sectional | An online survey fielded in June 2020 collected retrospective self-reports on everyday activities, stressors, and well-being of parents and caregivers at three timepoints: before local transmission, after local transmission but before "circuit breaker", and during "circuit breaker". | Results showed significant changes in jobs and income, childcare arrangement, family dynamics, and parents' emotional well-being throughout the three timepoints. Both mothers and fathers reported to worry most about the health and safety of family members and self. Mothers' time spent on housework partially mediated the effect of lockdown on their emotional well-being, and parents' conflict with other adults in the household partially mediated the effects of both pandemic and lockdown on their emotional well-being. The effects of pandemic and lockdown were also moderated by parents' age, education level, and fathers' authoritarian values. |
| **Zamarro, G., and M. Prados.** Gender Differences in Couples' Division of Childcare, Work and Mental Health During COVID-19. | Jul-20 | USA | *N* = 3980 adults, including parents of school-age children or children in daycare; 26,052 observations | Longitudinal | Uses data from eight waves of the online *Understanding Coronavirus in America Tracking Survey*, collected approximately every two weeks from March 10 to July 22, 2020 to understand gender differences within households on the impacts of COVID-19 on childcare provision, employment, working arrangements, and psychological distress levels. | Combining a measure of feelings of anxiety and depression, we find that psychological distress increased significantly early during the crisis, with 49% of mothers showing at least mild symptoms of psychological distress in early April, as compared to 41% of women without school-age children. (In the U.S., schools closed in all states between March 16-24) This new gap in psychological distress observed between mothers and women without school-age children appears to be driven by higher levels of psychological distress among mothers of elementary school-age and younger children. |
| **Zanardo, V., et al.** Psychological Impact of COVID-19 Quarantine Measures in Northeastern Italy on Mothers in the Immediate Postpartum Period. | Jun-20 | Italy | *N* = 192 mothers post-partum, specifically mothers of singleton, healthy neonates delivered between March 8 and May 3 | Case-control | Case-control study of mothers who gave birth during a COVID-19 quarantine period between March 8 and May 3, 2020, with a matched comparison group of matched postpartum women who delivered in the same period in 2019. Participants completed the Edinburgh Postnatal Depression Scale | A case-control study of mothers who gave birth during a COVID-19 quarantine period between March 8 and May 3, 2020 had significantly higher mean EPDS scores compared with the control group who delivered in the same period in 2019 (8.5 vs 6.34). |
|  |  |  |  |  | (EPDS) on the second day postpartum. |  |

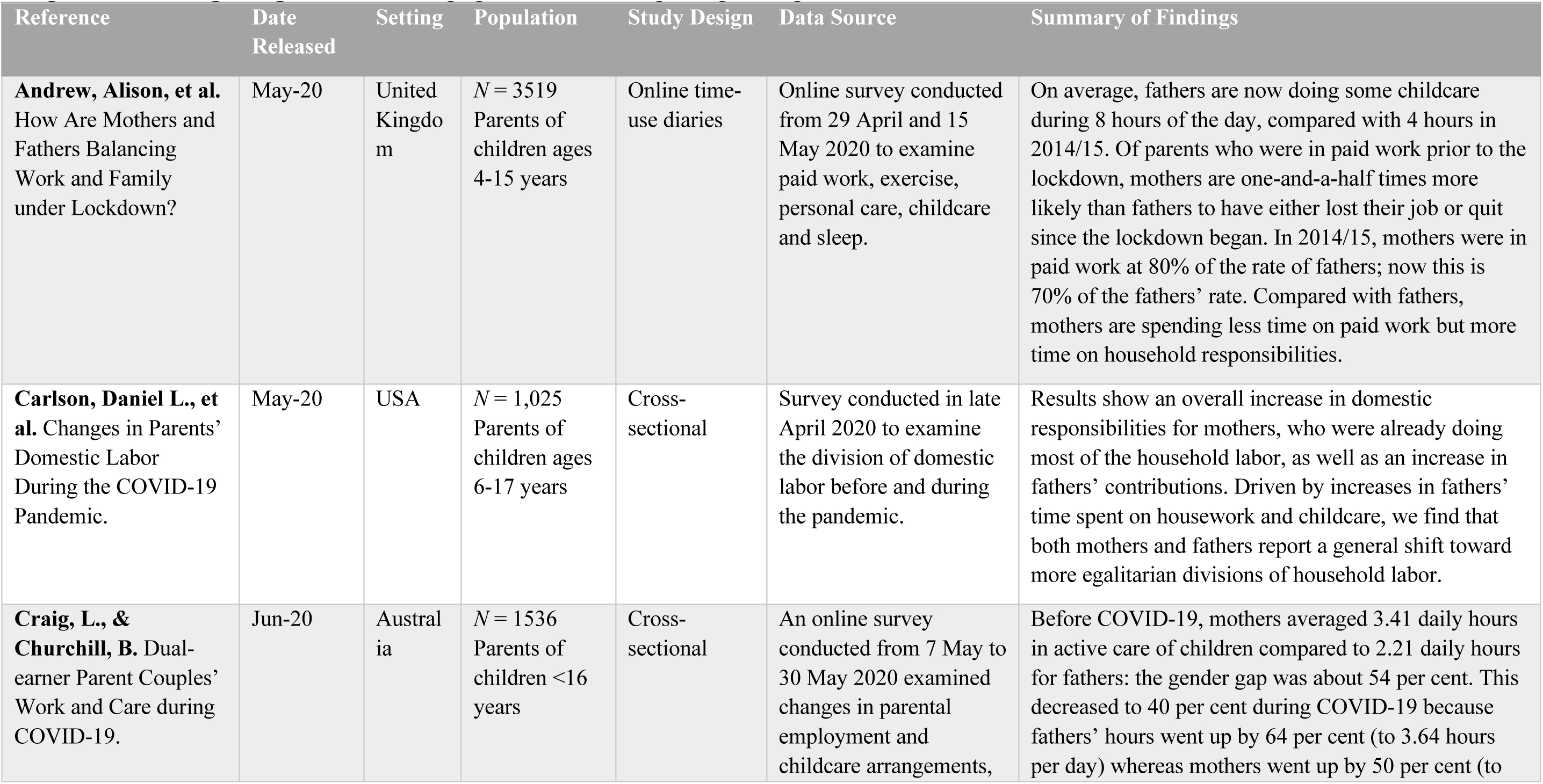
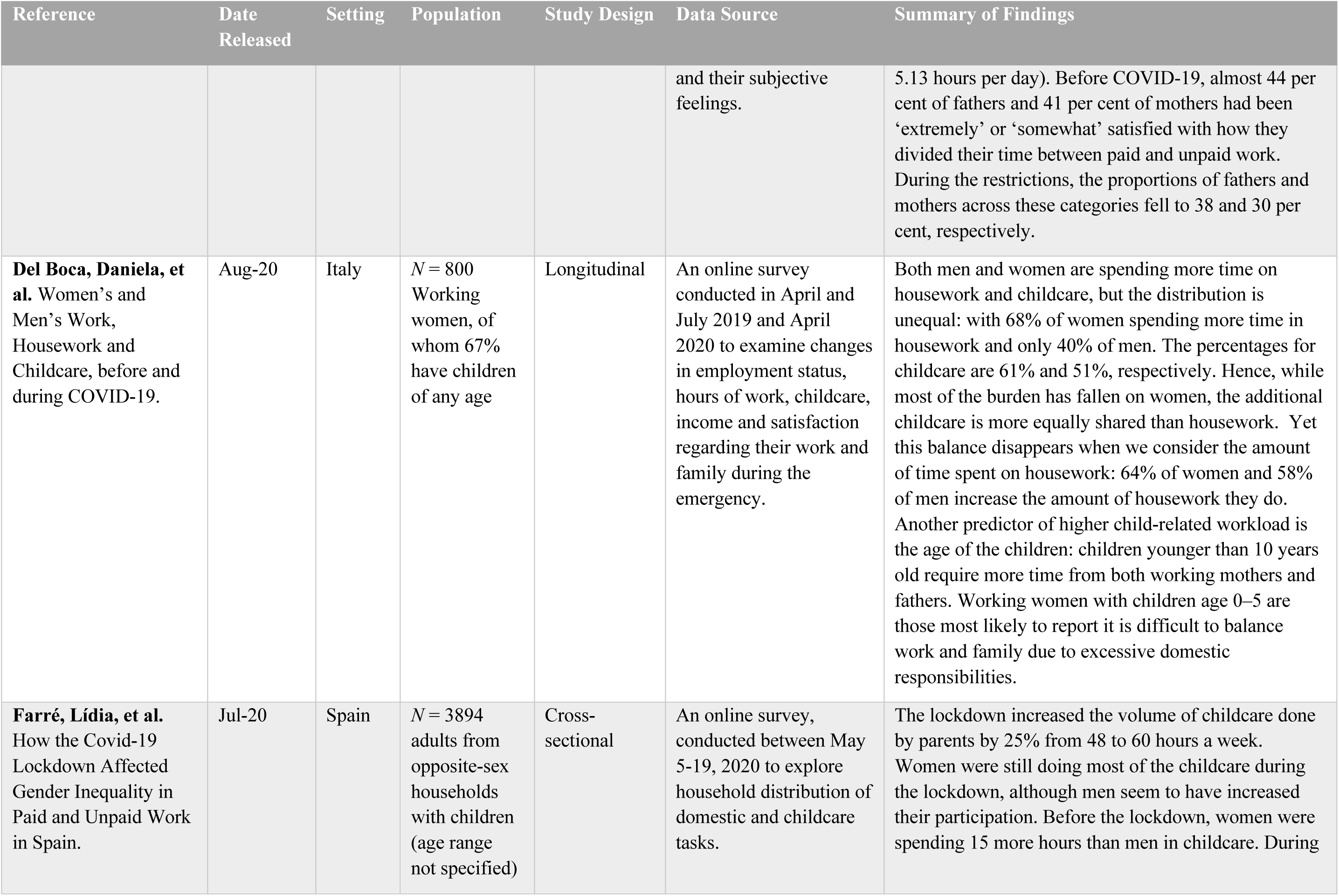
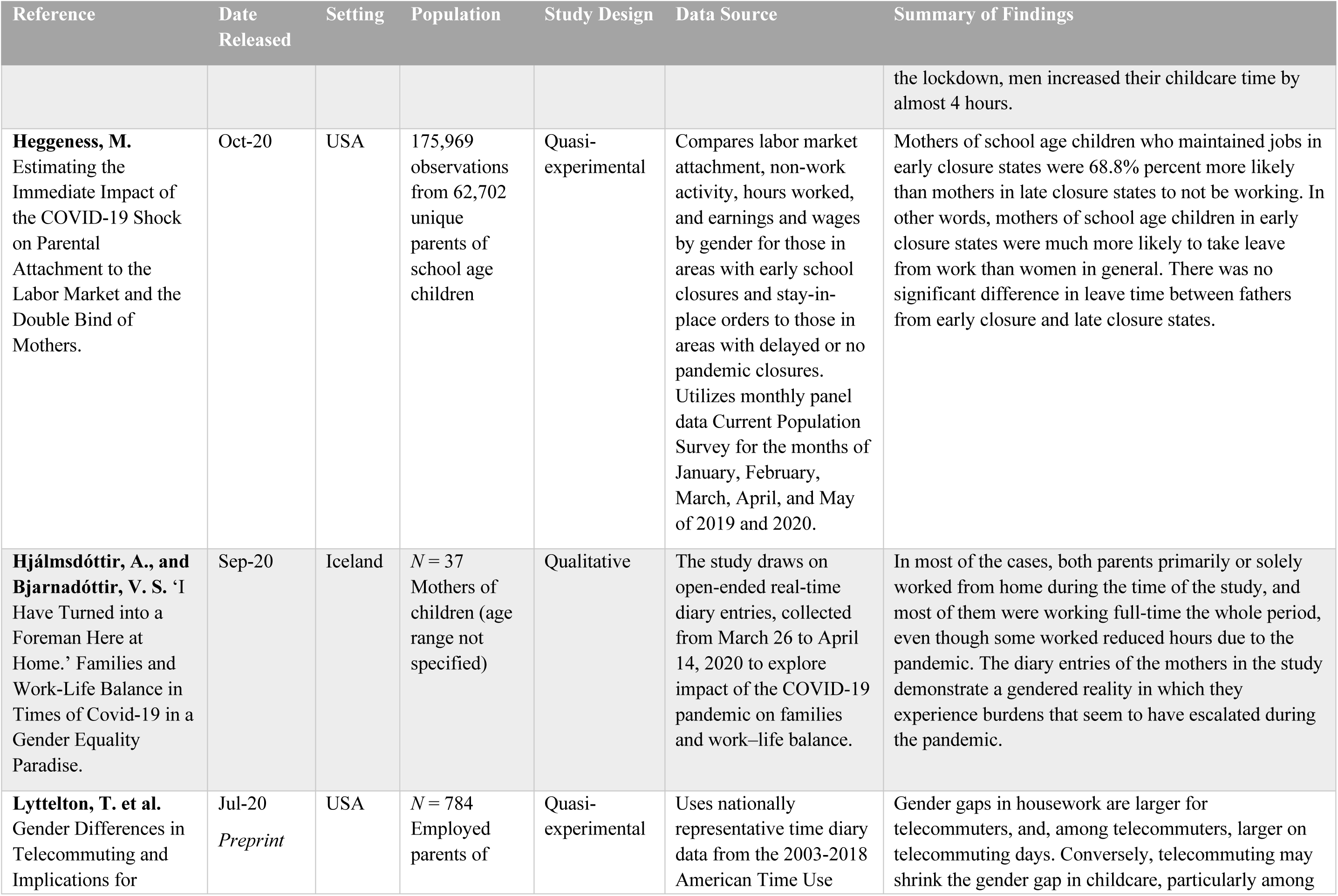
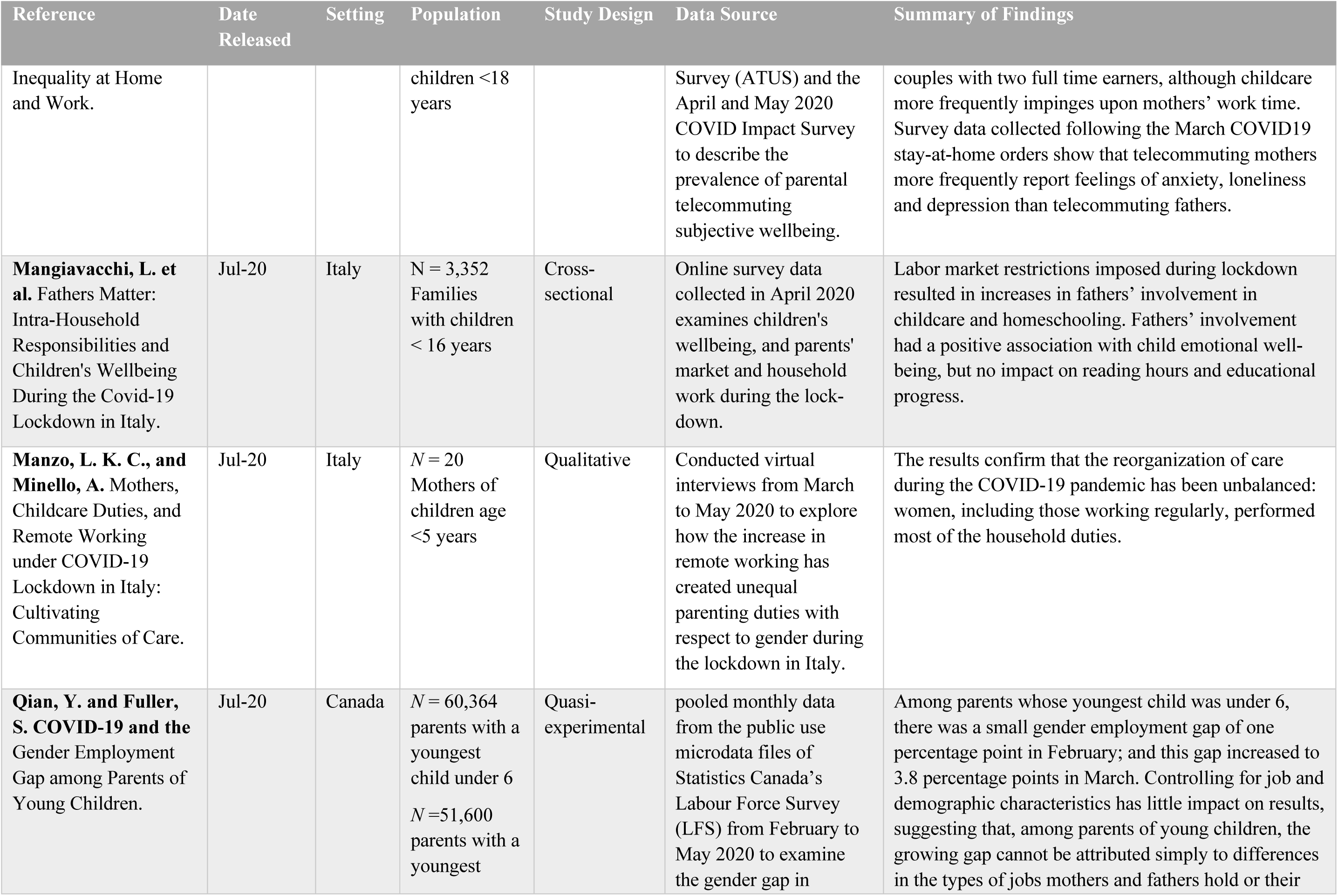
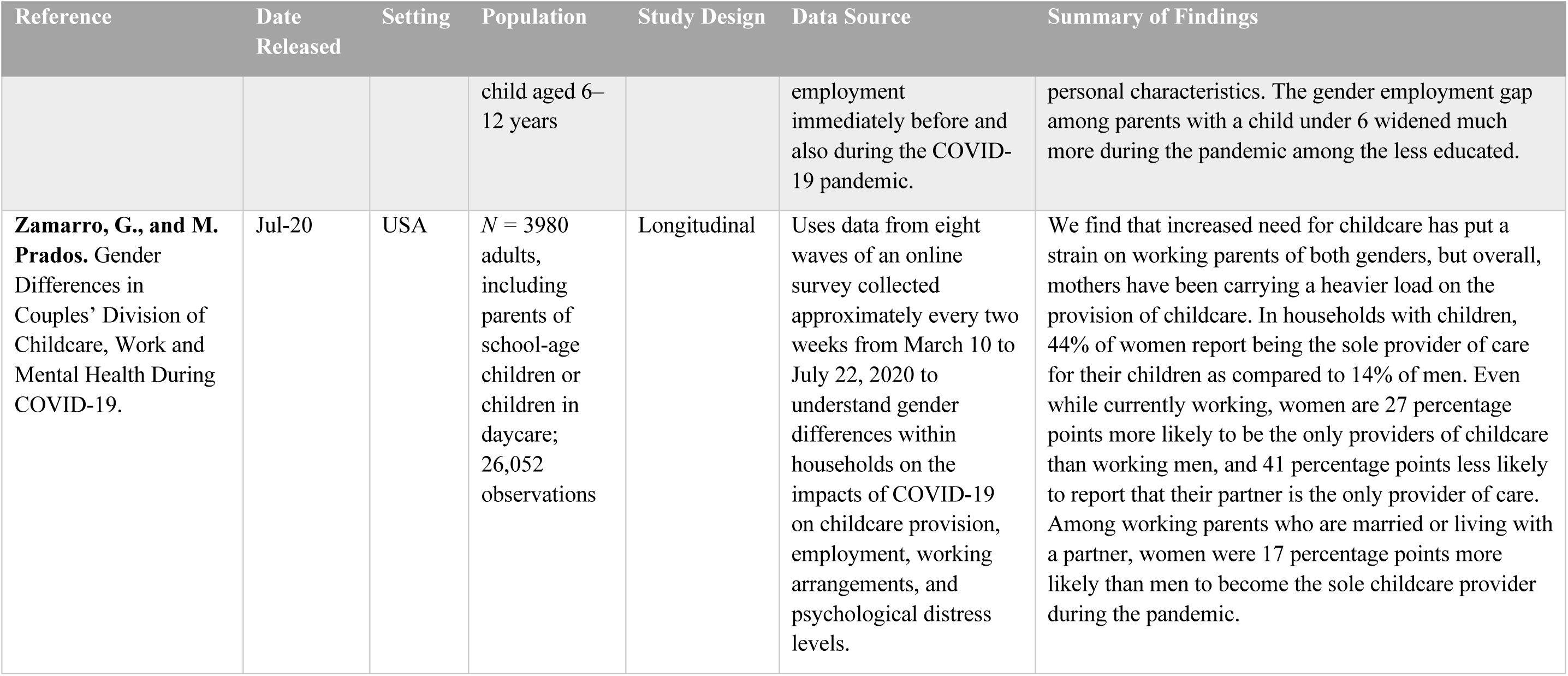
**Responsive caregiving:** Fathers’ engagement in caregiving and gendered division of labour

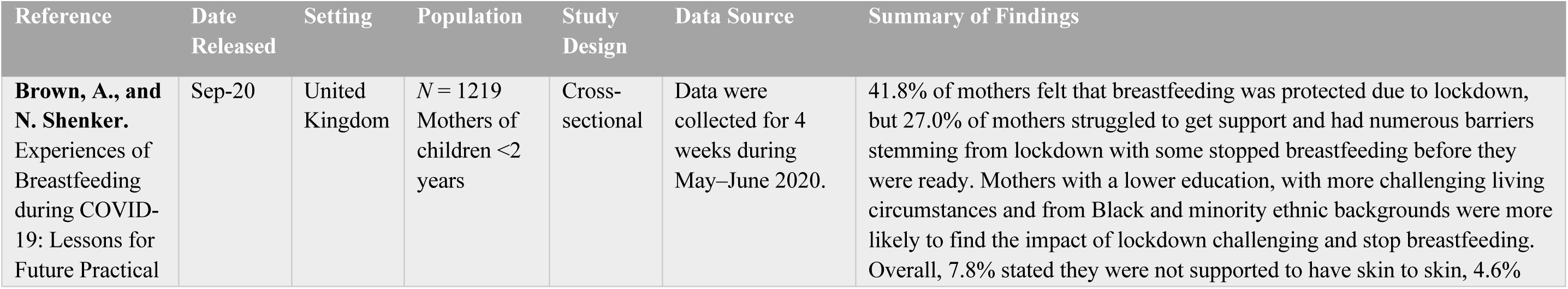
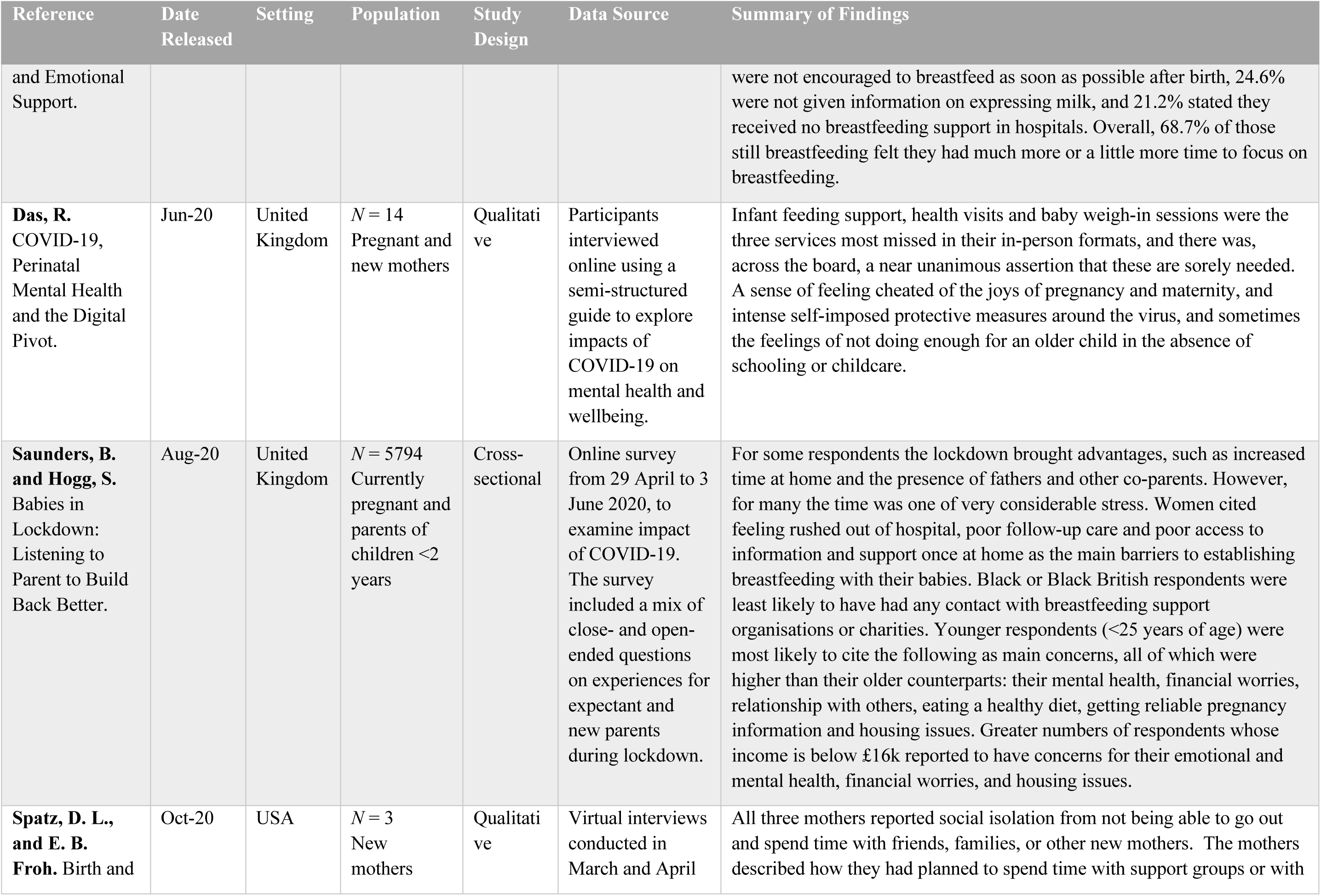
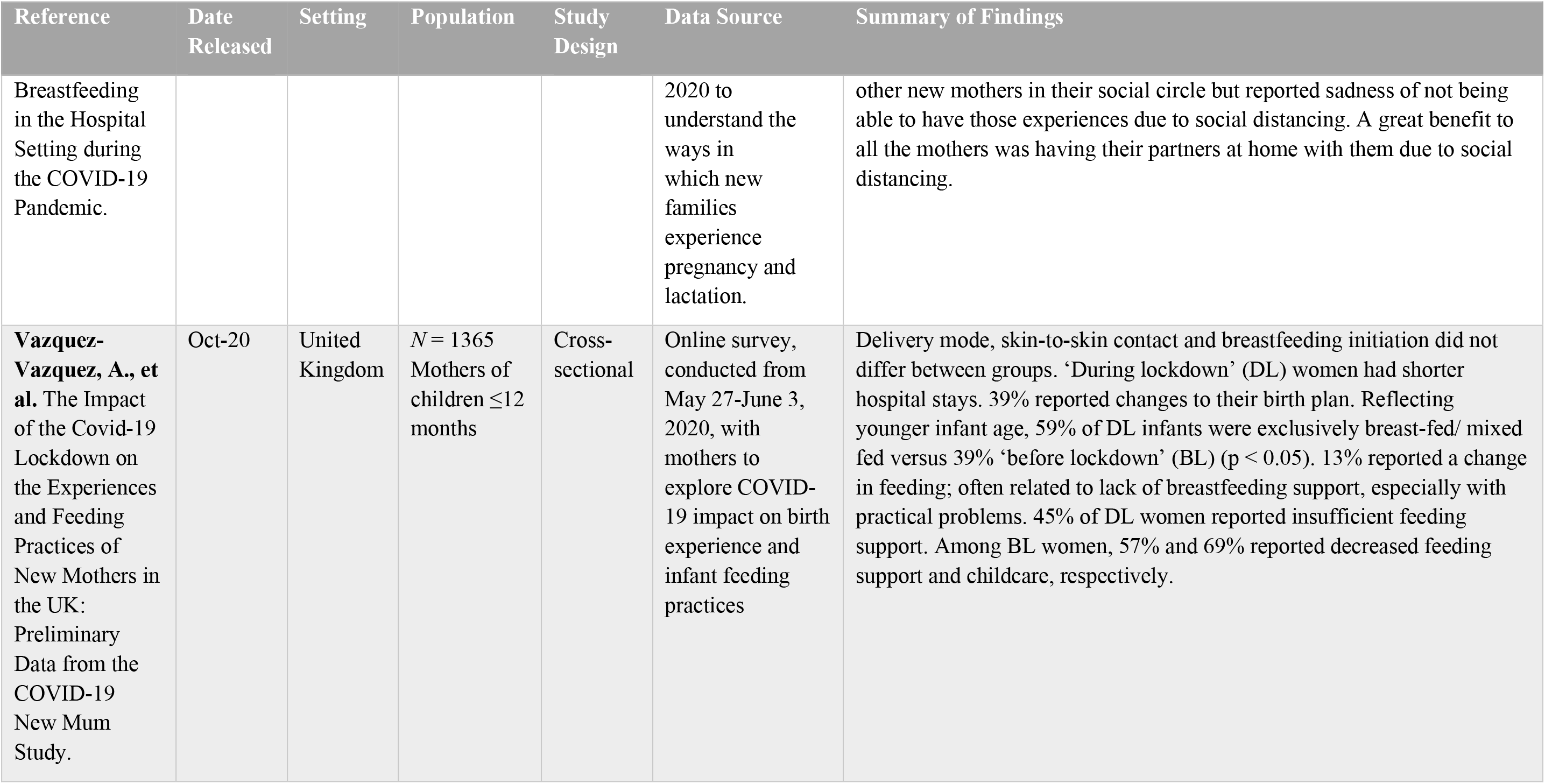
**Responsive caregiving:** breastfeeding support

Responsive caregiving: breastfeeding support
| Reference | Date Released | Setting | Population | Study Design | Data Source | Summary of Findings |
| --- | --- | --- | --- | --- | --- | --- |
| **Brown, A., and N. Shenker.** Experiences of Breastfeeding during COVID-19: Lessons for Future Practical | Sep-20 | United Kingdom | *N* = 1219 Mothers of children <2 years | Cross-sectional | Data were collected for 4 weeks during May–June 2020. | 41.8% of mothers felt that breastfeeding was protected due to lockdown, but 27.0% of mothers struggled to get support and had numerous barriers stemming from lockdown with some stopped breastfeeding before they were ready. Mothers with a lower education, with more challenging living circumstances and from Black and minority ethnic backgrounds were more likely to find the impact of lockdown challenging and stop breastfeeding. Overall, 7.8% stated they were not supported to have skin to skin, 4.6% |
| and Emotional Support. |  |  |  |  |  | were not encouraged to breastfeed as soon as possible after birth, 24.6% were not given information on expressing milk, and 21.2% stated they received no breastfeeding support in hospitals. Overall, 68.7% of those still breastfeeding felt they had much more or a little more time to focus on breastfeeding. |
| **Das, R.**<br>COVID-19, Perinatal Mental Health and the Digital Pivot. | Jun-20 | United Kingdom | *N* = 14<br>Pregnant and new mothers | Qualitative | Participants interviewed online using a semi-structured guide to explore impacts of COVID-19 on mental health and wellbeing. | Infant feeding support, health visits and baby weigh-in sessions were the three services most missed in their in-person formats, and there was, across the board, a near unanimous assertion that these are sorely needed. A sense of feeling cheated of the joys of pregnancy and maternity, and intense self-imposed protective measures around the virus, and sometimes the feelings of not doing enough for an older child in the absence of schooling or childcare. |
| **Saunders, B. and Hogg, S.**<br>Babies in Lockdown: Listening to Parent to Build Back Better. | Aug-20 | United Kingdom | *N* = 5794<br>Currently pregnant and parents of children <2 years | Cross-sectional | Online survey from 29 April to 3 June 2020, to examine impact of COVID-19. The survey included a mix of close- and open-ended questions on experiences for expectant and new parents during lockdown. | For some respondents the lockdown brought advantages, such as increased time at home and the presence of fathers and other co-parents. However, for many the time was one of very considerable stress. Women cited feeling rushed out of hospital, poor follow-up care and poor access to information and support once at home as the main barriers to establishing breastfeeding with their babies. Black or Black British respondents were least likely to have had any contact with breastfeeding support organisations or charities. Younger respondents (<25 years of age) were most likely to cite the following as main concerns, all of which were higher than their older counterparts: their mental health, financial worries, relationship with others, eating a healthy diet, getting reliable pregnancy information and housing issues. Greater numbers of respondents whose income is below £16k reported to have concerns for their emotional and mental health, financial worries, and housing issues. |
| **Spatz, D. L., and E. B. Froh.** Birth and | Oct-20 | USA | *N* = 3<br>New mothers | Qualitative | Virtual interviews conducted in March and April | All three mothers reported social isolation from not being able to go out and spend time with friends, families, or other new mothers. The mothers described how they had planned to spend time with support groups or with |
| Breastfeeding in the Hospital Setting during the COVID-19 Pandemic. |  |  |  |  | 2020 to understand the ways in which new families experience pregnancy and lactation. | other new mothers in their social circle but reported sadness of not being able to have those experiences due to social distancing. A great benefit to all the mothers was having their partners at home with them due to social distancing. |
| **Vazquez-Vazquez, A., et al.** The Impact of the Covid-19 Lockdown on the Experiences and Feeding Practices of New Mothers in the UK: Preliminary Data from the COVID-19 New Mum Study. | Oct-20 | United Kingdom | *N* = 1365 Mothers of children ≤12 months | Cross-sectional | Online survey, conducted from May 27-June 3, 2020, with mothers to explore COVID-19 impact on birth experience and infant feeding practices | Delivery mode, skin-to-skin contact and breastfeeding initiation did not differ between groups. ‘During lockdown’ (DL) women had shorter hospital stays. 39% reported changes to their birth plan. Reflecting younger infant age, 59% of DL infants were exclusively breast-fed/ mixed fed versus 39% ‘before lockdown’ (BL) (*p* < 0.05). 13% reported a change in feeding; often related to lack of breastfeeding support, especially with practical problems. 45% of DL women reported insufficient feeding support. Among BL women, 57% and 69% reported decreased feeding support and childcare, respectively. |

**Opportunities for early learning**

Opportunities for early learning
| Reference | Date Released | Setting | Population | Study Design | Data Source | Summary of Findings |
| --- | --- | --- | --- | --- | --- | --- |
| **Barnett, S., Jung, K., & Nores, M.** Young children's home learning and preschool | Aug-20 | USA | *N* = 945 Parents of children ages 3-5 years | Cross-sectional | The survey was conducted between May 22 and June 5, 2020 about child's | The 2020 NIEER survey reported frequencies of engaging in reading, singing, teaching activities, and arts and crafts were lower than those in the 2019 NHES. The percentage reporting storytelling three or more times in the past week was higher than before the pandemic. Nearly three-quarters (74%) of |
| participation experiences during the pandemic.<br>NIEER 2020<br>Preschool Learning Activities Survey |  |  |  |  | daily activities during the pandemic. | children attending preschool programs had their programs closed as a result of the pandemic. More than 90% of public program classrooms closed compared to two-thirds of private programs. |
| **Briggs, D. C.**<br>COVID-19: The Effect of Lockdown on Children's Remote Learning Experience – Parents' Perspective. | Oct-20 | Nigeria | *N* = 260<br>parents of schoolchildren | Cross-sectional | An online survey distributed from May 9-June 8, 2020 explored parents' perspectives on education during stay-at-home periods | 77.7% possess a computer device, 93.2% have internet services and 55.0% spend between N5,000 – N9,999 (13.1 – 26.2 USD) per month for internet services. Only 35.4% had their children participate in the ongoing radio/TV learning sessions, 46.1% educate their children with their computer devices at home, however, 40.0% preferred an online based method of homeschooling. Overall, 70.0% were dissatisfied with the level of homeschooling their children received. Respondents with higher education and income level were significantly more likely to possess a computer device, have internet access at home and prefer online classes when compared to low-income earning parents with secondary education and below. |
| **Dayal, H. C., & Tiko, L.** When are we going to have the real school? A case study of early childhood education and care teachers' experiences surrounding education during the | Oct-20 | Small Pacific Island | *N* = 4<br>teachers/staff at early childhood education and care centres | Qualitative | Telephone interview were conducted to allow teachers to share their feelings and emotions about the pandemic and school closures; plans for delivering education during the pandemic; and proposed strategies | Teachers worried about students' health and safety and general well-being during complete lockdown. In one facility, each teacher created a Viber group with parents (a free app that has text, calling and video-calling features) to communicate about general well-being of children and to send learning materials. Parents who cannot access Viber are called by teachers on their phones. Teachers mentioned the need for more training in technical issues. Another facility is using Zoom for continuing education. Staff noted approximately 80% of students were able to continue education, including children who had returned to Australia and New Zealand. Staff had a single laptop and modem, so teachers took turns |
| COVID-19 pandemic. |  |  |  |  | to overcome challenges. | delivering their virtual lessons at different time. Some parents did not have Zoom enabling resources at home or were unaware of how to use them. The latter was mainly for children who were looked after by grandparents, who may have lacked knowledge to use online technologies. Parents who faced such difficulties had 'print copy' teaching materials sent to them through email, or in a few cases, delivered to their homes by staff. |
| **Dong, C., et al.**<br>Young Children's Online Learning during COVID-19 Pandemic: Chinese Parents' Beliefs and Attitudes. | Sep-20 | China | *N* = 3275<br>Parents of children enrolled in ECE | Cross-sectional | This survey study was to explore parental beliefs and attitudes about online learning. | Most parents (92.7%) reported that their children had online learning experiences during the pandemic, and many (84.6%) spent less than a half-hour each time. The parents generally had negative beliefs about the values and benefits of online learning and preferred traditional learning in early childhood settings. They tended to resist and even reject online learning for three key reasons: shortcomings of online learning, young children's inadequate self-regulation, and their lack of time and professional knowledge in supporting children's online learning. Also, the hardship caused by the COVID-19 pandemic has made them suffer, thus more resistant to online learning at home. |
| **Grover, S., et al.** A Survey of Parents of Children Attending the Online Classes During the Ongoing COVID-19 Pandemic. | Oct-20 | India | *N* = 289<br>guardians of children attending online classes | Cross-sectional | An online survey carried out between June 21 and July 17, 2020 assessed the impact of online classes on children and their parents or guardians. | A majority (93.8%) of children were studying in private schools and attending online live classes (83.7%). About three-fourth of the parents/guardians reported that online classes were less comfortable (81.7%) and less satisfactory (78.5%); guardians reported that children had poor attention and concentration (80.6%), lower learning of theoretical and practical aspects of the subject (82.4%), and lower ability to sit satisfactorily for the duration of the class (78.6%). The most common reported distraction was surfing internet (52.2%) or online games (52.2%). |
| **Kim, J., et al.** The Implications of COVID-19 on Early Learning Continuity in Ethiopia: Perspectives of Parents and Caregivers. | Oct-20 | Ethiopia | *N* = 480<br>Parents of children <5 | Cross-sectional | mobile phone surveys to understand how parents and caregivers with pre-primary aged children have | More than half of respondents reported they do not have children's books or learning materials at home, especially caregivers who are not literate. A large difference between urban and rural locations was also observed for families' access to electricity and technologies to support children's learning. Most have received little support from schools or local governments. Only 10% of caregivers reported they have been in contact with pre-primary teachers or school principals, with significant differences by household wealth and across regions. Only half report they have engaged in supporting learning activities for pre-primary children, which favours families living in urban areas. Mothers are most likely to be responsible for supporting children's learning at home; yet mothers are less likely to be literate. Around three quarters reported they play more often with their child since the COVID-19 crisis than before, including telling stories or singing songs. However, caregivers from poorer households or who are illiterate are less likely to engage in such activities. Only 12% said they had used radio lessons with their children. |
| **Lee, S. J., et al.** Parenting Activities and the Transition to Home-Based Education During the COVID-19 Pandemic. | Oct-20<br>*Preprint* | USA | *N* = 405<br>Parents with children <12 years | Mixed methods | An online survey, including open-ended questions, conducted from April 24-25, 2020, examined parental mental health and the transition to home-based learning. | 78% of parents said they were educating their child at home due to COVID19. Most (77.1%) reported use of online tools for at-home education, including educational apps, social media, and school-provided electronic resources. Most parents were spending more time involved in daily caregiving of their children since COVID-19. Content analyses of open-ended questions indicated that school closures were a significant disruption, followed by lack of physical activity, and social isolation. |
| **Mangiavacchi, L. et al.** Fathers Matter: Intra-Household | Jul-20 | Italy | *N* = 3,352<br>families | Cross-sectional | Online survey data collected in April 2020 examines | Only about 60% of primary school children had some live online classes and about 30% received only some learning material accompanied by homework to be delivered through |
| Responsibilities and Children's Wellbeing During the Covid-19 Lockdown in Italy. |  |  | with children < 16 years |  | children's wellbeing, and parents' market and household work during the lock-down. | an electronic registry. Only 18% of children aged 3-5 had some live on-line classes, and 37% received some material from their teachers. 45% of children aged 3-5 were never involved in any school activity at the date of interview. Changes in reading to preschool children, who listen to their parents reading, increased by almost half an hour more per day. During lockdown, all children watched about twice as much TV (about 3 hours for kindergarten and primary school children). Labor market restrictions imposed during lockdown resulted in increases in fathers' involvement in childcare and homeschooling. Fathers' involvement had a positive association with child emotional well-being, but no impact on reading hours and educational progress. |
| **World Vision.**<br>Rapid Assessment of the Impact of COVID-19 on Child Wellbeing in Cambodia. | Jun-20 | Cambodia | N = 222 household heads (survey); 42 key informants (local leaders, etc.); 65 Agricultural Cooperative leaders (survey) | Mixed methods | The rapid assessment was conducted from May 11-14, 2020 to assess food insecurity; education; and livelihoods among vulnerable families. | Confirmed that home learning is very challenging for children for a variety of reasons, including limited availability of learning materials at home and limited support received by children. Technological barriers (bad internet connectivity, costs of internet connection and limited availability of a device to connect) while significant were not the highest reported challenges. Overall, there was a consensus that school closures are going to lead to school dropout, which respondents link to the impact that COVID-19 is having on livelihoods and to the perceived increase in child labour. |

**Opportunities for early learning:** Outdoor play and physical activity

Opportunities for early learning: Outdoor play and physical activity
| Reference | Date released | Setting | Population | Study Design | Data Source | Summary of Findings |
| --- | --- | --- | --- | --- | --- | --- |
| **Carroll, N., et al.**<br>The Impact of COVID-19 on Health Behavior, | Aug-20 | Canada | N = 254 Parents of children ages 18 | Cross-sectional | An online survey administered between 20 April 2020 to 15 May 2020 to explore impact of COVID-19 on physical | Many parents expressed concern about decreases in frequency and intensity of children's physical activity and increases in screen time. Parents reported physical activity is limited due to lack of space and variety in tools and toys. |
| Stress, Financial and Food Security among Middle to High Income Canadian Families with Young Children. |  |  | months to 5 years |  | activity, screen time, and sleep among parents and children, as well as stress, financial and food insecurity of families. | Parents also reported strategies used to increase physical activity, such as scheduling outdoor playtime. Specific resources that were frequently mentioned included resources on engaging children in physical activity or any other activities, e.g., crafts, cooking to decrease screen time, tips for grocery shopping during COVID-19, homeschooling, scheduling, and time management. |
| **de Lannoy, L. et al.** Regional Differences in Access to the Outdoors and Outdoor Play of Canadian Children and Youth during the COVID-19 Outbreak. | Oct-20 | Canada | *N* = 1472<br>Parents of children ages 5–17 years | Cross-sectional | Used online survey data collected in April 2020. Parents were asked about their child's physical activity, sleep, sedentary time, time spent outdoors and in outdoor play in the last week and were asked to compare behaviours before and during the COVID-19 outbreak. | Change in child and youth outdoor behaviour ranged from 1 (much less play) to 5 (much more play) with 3 representing no change. Overall, we found that all regions exhibited a decrease in time spent outdoors and in outdoor play (i.e., below 3), with Ontario having experienced the greatest decline in both time spent outdoors and in outdoor play ($p < 0.001$). |
| **Dodd, H., et al.** Co-SPYCE Report One: Findings from 1728 parents/carers of 2-4 year olds on stress, child activities, child worries and need for support. | May-20 | UK | *N* = 1728<br>caregivers of children aged 2-4 years | Cross-sectional | An online survey was completed between April 17 April and May 20, 2020 to explore parent/carer stress, how children are spending time, parent feelings about child play, and the need for parental support. | Over 80% of children are reported by participating parents/carers to be getting at least 30 minutes of exercise a day but only 22% are reported to be getting the recommended 3 hours. Almost half of the children (47%) are reported to be spending no time playing with another child in their household. The top 3 stressors for participating parents/carers were (i) work, (ii) child's screen time, (iii) their child's wellbeing. Participating parents/carers particularly want support around managing children's emotions, educational demands, behaviours and coming out of social isolation. |
| **Drouin, M., et al.** How Parents and Their Children Used Social Media and Technology at the Beginning of the COVID-19 | Jul-20 | USA | *N* = 260<br>Parents of children ≤18 months | Cross-sectional | Online survey data, collected from March 20-25, 2020, explored children's use of social media and technology during the COVID-19 pandemic and its association with anxiety. | Mean differences revealed that children in the 0 to 5 year age range did not increase their technology use as much as older age groups, nor did they use social media/ social technology or video/phone calls as frequently. |
| Pandemic and Associations with Anxiety. |  |  |  |  |  |  |
| **Dunton, G., Do, B., & Wang, S.** Early Effects of the COVID-19 Pandemic on Physical Activity and Sedentary Behavior in US Children. | Sep-20 | USA | *N* = 211<br>Parents and guardians of children ages 5-13 years | Cross-sectional | online survey between April 25–May 16, 2020 assessed physical activity and sedentary behavior in previous day, as well as perceived changes in activities between pre-COVID-19 period and early-COVID-19 periods. | The most common physical activities during the early-COVID-19 period were free play/unstructured activity (e.g., running around, tag) (90% of children) and going for a walk (55% of children). Children engaged in about 90 min of school-related sitting and over 8 h of leisure-related sitting a day. Girls and older children generally spent more time in sedentary behaviors than boys and younger children. Children were more likely to perform physical activity at home indoors or on neighborhood streets during the early- vs. pre-COVID-19 periods. |
| **Guerrero, M. D., et al.** Canadian Children's and Youth's Adherence to the 24-h Movement Guidelines during the COVID-19 Pandemic: A Decision Tree Analysis. | Jun-20 | Canada | *N* = 1472<br>Parents of children ages 5-17 years | Cross-sectional | Online survey fielded in April 2020 explored adherence and non-adherence to Canadian 24-hr movement guidelines during the COVID-19 outbreak. Survey assessed demographic, social, and environmental characteristics, as well as physical activity, screen time, and sleep. | Characteristics associated with adherence to recommendations included high parental perceived capability to restrict screen time, annual household income of $\geq \$100,000$, increases in children's and youth's outdoor physical activity/sport since the outbreak began, being a boy, having parents younger than 43 years old, and small increases in children's and youth's sleep duration since the outbreak began. Characteristics associated with non-adherence to the recommendations included low parental perceived capability to restrict screen time, youth aged 12-17 years, decreases in children's and youth's outdoor physical activity/sport since the outbreak began, primary residences located in all provinces except Quebec, low parental perceived capability to support children's and youth's sleep and physical activity, and annual household income of $\leq \$99,999$. |
| **Mitra, R., et al.** Healthy Movement Behaviours in Children and Youth During the COVID-19 | Aug-20 | Canada | *N* = 1472<br>Parents of children ages 5-17 years | Cross-sectional | Secondary analysis of an online survey conducted in April 2020 explores patterns of physical activity, sedentary and sleep behaviours among children and youth during the COVID-19 pandemic. The | Cluster analysis identified two groups (those with increased outdoor activities, those with decreased outdoor activities). Compliance to 24-hour movement guidelines was low among both groups. For children (aged 5-11), living in houses (versus apartments) was correlated with increased outdoor activities; proximity to major roads was a barrier. Similarly, dwelling density was negatively associated with |
| Pandemic: Exploring the Role of the Neighbourhood Environment. |  |  |  |  | survey included health behavior measures and built environment measures. | increased outdoor activity, suggesting that those living in high-density neighbourhoods may demonstrate less healthy movement behaviours during a pandemic. However, results also indicate that for those living in higher density areas, access to parks within 1 km increased the probability of being in cluster 1 (i.e. increased outdoor activities). |
| **Moore, S. A., et al.** Impact of the COVID-19 Virus Outbreak on Movement and Play Behaviours of Canadian Children and Youth: A National Survey. | Jul-20 | Canada | *N* = 1472 Parents of children ages 5–17 years | Cross-sectional | Online survey assessed immediate changes in child movement and play behaviours during the COVID-19 outbreak. Behaviours included physical activity and play, sedentary behavior, and sleep. | Only 4.8% (2.8% girls, 6.5% boys) of children and 0.6% (0.8% girls, 0.5% boys) of youth were meeting combined movement behaviour guidelines during COVID-19 restrictions. Children and youth had lower PA levels, less outside time, higher SB (including leisure screen time), and more sleep during the outbreak. Parental encouragement and support, parental engagement in PA, and family dog ownership were positively associated with healthy movement behaviours. Although families spent less time in PA and more time in SB, several parents reported adopting new hobbies or accessing new resources. |
| **Xiang, M., et al.** Impact of COVID-19 Pandemic on Children and Adolescents' Lifestyle Behavior Larger than Expected. | Jul-20 | China | *N* = 2426 children and adolescents (6–17 years) | Quasi-experimental | Longitudinal online surveys conducted from 3-21 January and 13-23 March 2020 to better understand physical activity and sedentary behavior among school children. | Overall, the median time spent in physical activity decreased drastically, from 540 min/week before the pandemic to 105 min/week during the pandemic. During the pandemic, prevalence of physically inactive students increased from 21.3% to 65.6%. Screen time considerably increased during the pandemic in total (+1730 min [or approximately 30 h] per week on average). Screen time during leisure was also prolonged, indicating that nearly a quarter of students engaged in long screen time for leisure. |

**Safety and Security:** Child protection from violence and neglect

Safety and Security: Child protection from violence and neglect
| Reference | Date Released | Setting | Population | Study Design | Data Source | Summary of Findings |
| --- | --- | --- | --- | --- | --- | --- |
| **Barboza, G. E., et al.** A Spatiotemporal Analysis of the Impact of COVID-19 on Child Abuse and Neglect in | Sep-20 | USA | Children <18 years | Quasi-experimental | Utilizes police reports from before and during the COVID-19 pandemic (July 24, 2019–January 20, 2020 vs. January 21–July 19, 2020) to explore association between implementation of social distancing protocols and | Results indicate a significant 8% decrease in the number of child abuse and neglect reports during the COVID-19 pandemic (January 21, 2020–July 19, 2020) compared to the time period immediately preceding it (July 24, 2019–January 20, 2020). Spatiotemporal analysis identified locations of |
| the City of Los Angeles, California. |  |  |  |  | reported child abuse and neglect. Neighborhood structural factors (e.g., school absenteeism, poverty, unemployment, housing insecurity and birth assets) are explored. | emerging "hotspots" for reported child abuse and neglect during the pandemic. Neighbourhoods characterized by severe housing burden, the number of assets children have at birth, poverty, school absenteeism and labor force participation were significantly associated with new and intensifying hotspots of child abuse and neglect. |
| **Baron, E. Jason, et al.**<br>Suffering in Silence: How COVID-19 School Closures Inhibit the Reporting of Child Maltreatment. | Aug-20 | USA | School age children | Quasi-experimental | Uses publicly available, monthly county-level data for Florida from January 2004 to April 2020 to estimate the effect of COVID-19 school closures on the number of child maltreatment allegations. Leverage monthly data to predict the number of allegations that would be expected due to seasonal fluctuations and secular trends in March and April 2020. | Using counterfactual distribution of county-level data from Florida, results show the number of reported child maltreatment allegations was approximately 27% or 15,000 lower than expected for the months of March and April 2020, the first two months in which schools closed. Counties with higher numbers of educators trained to identify and report child maltreatment (e.g., school psychologists and school nurses) experience a disproportionately larger reduction in the number of child maltreatment allegations. |
| **Bhopal, S., et al.** Who Has Been Missed? Dramatic Decrease in Numbers of Children Seen for Child Protection Assessments during the Pandemic. | Jun-20 | United Kingdom | Children ≤16 years | Quasi-experimental | Examined monthly referral data for children and young people attending child protection medical examination as part of a child safeguarding assessment from January to April 2020 in comparison with the same months in the last 2 years. | Results showed a one-third reduction in child protection assessments from January to April 2020, compared to previous years. The total number of assessments from January to April was 152 in 2018, 156 in 2019 and 99 in 2020. |
| **Branstetter, S., et al.**<br>The Impact of the COVID-19 Pandemic on Families in Germany. | Oct-20<br>*Preprint* | Germany | N = 918 parents of children ages 18 months to 6 years | Cross-sectional | Online survey assessing the impact of the COVID-19 pandemic on families with young children from areas with low and moderate COVID-19 prevalence were fielded at the end of the German lock-down period. Outcomes were parent-reported impact of the COVID-19 pandemic on family life, concerns and trust in | Many participants reported an impact of the COVID-19 pandemic on social and family life. About one third of the families (30.5%) were at least very much burdened by the situation, and the lack of social contacts was very/extremely bothersome for almost half of families. On the other hand, the majority coped well with the situation. About half had a clear daily schedule. Families indicated that they were enjoying the additional time they could spend together (60%) and that they had united as a family (56%). Nevertheless, 22.5% |
|  |  |  |  |  | political measures. Participants were part of two ongoing cohort studies. | of the families reported relevantly more conflicts with children and 19% and 12% reported more conflicts between children or between the spouses, respectively. |
| **Bullinger, L., et al.** COVID-19 and Alleged Child Maltreatment. | Oct-20 | USA | Children <18 years | Quasi-experimental | Compared weekly, county-level referral data from January 8, 2018 through May 30, 2020 for all 159 counties in Georgia to examine whether COVID-19 related policies affected child maltreatment referrals. Superimposed aggregate, anonymous cell phone tracking data to determine whether time at home is related to referrals. | Relative to 2018 and 2019 trends during the same time period, referral rates plummeted by approximately 55% following the emergency declaration. Maltreatment referrals begin to slowly return to pre-COVID-19 levels around 7 weeks following the emergency declaration, but never fully rebound during the study period. As time at home during the pandemic increased, the risk of supervisory neglect increased, with largest impacts in metropolitan counties and counties with more poverty. |
| **Bullinger, L. R., et al** COVID-19 and Crime: Effects of Stay-at-Home Orders on Domestic Violence. | Jul-20 | USA | Children <18 years | Quasi-experimental | Uses cell phone block-level activity data and administrative 911 and crime data from 2019 and 2020 for Chicago to estimate the effects of Stay-at-Home Orders on calls for police service, crimes recorded by police, and arrests made relating to domestic violence. | Results show domestic-related police calls increased by 8% during the stay at home order. The effects are largest in areas with more households that are married with children, areas with relatively high rates of rental residences, and areas that experienced the largest changes in staying at home. Did not find significant effects for domestic crimes against children, as measured by physical child abuse. This result may indicate that although domestic crimes rose in March-April 2020, calls for child abuse fell, potentially due to lessened visibility of child victims and limitations of various reporting channels for child maltreatment crimes. |
| **Cabrera-Hernández, F., and Padilla-Romo, M.** Hidden Violence: How COVID-19 School Closures Reduced the Reporting of Child Maltreatment. | Jul-20 | Mexico | Children ages 3-15 years | Quasi-experimental | Leverage incident-level crime reports from Mexico City Attorney General's Office for January 2019-June 2020 to estimate the impact of school closures on the reporting of school-age child maltreatment. | The results show an average reduction of 21% and 30% in March 2020 for the reporting of child maltreatment among school-aged children, or a reduction of 7 to 10 cases from the average of 33 per week. These effects are higher for females, at a reduction of 66%, or 22 cases per week from an average of 33; and in poorer municipalities (reduction of 40% or 14 cases less a week, from an average of 34). These effects suggest that 220 incidents of child maltreatment for school-age females were not reported to the authorities in the 10 weeks after school closures. |
| **Chung, G., et al.** Mediating Effects of Parental Stress on Harsh Parenting and Parent-Child Relationship during Coronavirus (COVID-19) Pandemic in Singapore. | Jun-20 | Singapore | *N* = 258 Parents of children <12 years | Cross-sectional | An online survey disseminated from April 22, 2020, to May 5, 2020 explored work, family life and parenting during the pandemic. | Parents who were more impacted by COVID-19 experienced more parenting stress than those who felt they were less impacted by COVID-19, and parents who experienced more parenting stress indicated that they had used more harsh parenting and felt less close with their children in the past weeks. There was no evidence that parents' perceived impact of COVID-19 influenced harsh parenting or parent-child relationships independent of its effect on parenting stress. |
| **Dulieu, N. and Burgess, M.** The Hidden Impact of COVID-19 on Child Rights. | Sep-20 | Multiple countries (37 countries) | *N* = 17,565 Parents and caregivers of children <18 years | Cross sectional | An online survey assessed implications of COVID-19 pandemic on Child Rights. Measured household-level characteristics, child rights, caregiver's well-being and family relationships, child education and well-being, among other topics. Sample included Save the Children program participants, specific population groups of interest to the program, and the general public. | Nearly one-third (32%) of the households reported that violence had occurred in the home, including children and/or adults being verbally or physically abused. Parents/caregivers in urban locations reported a higher rate of violence in the home (23%) compared to rural respondents (17%). A statistically significant positive correlation was found between reported income loss and reported levels of household violence. |
| **Edwards, J.** Protect a Generation: The Impact of COVID-19 on Children's Lives. | Sep-20 | Multiple countries (16 countries) | *N* = 17,565 Parents and caregivers of children <18 years | Mixed methods | An online survey conducted with Save the Children participant households assessed implications of COVID-19 pandemic on child rights, caregiver's well-being and family relationships, child education and well-being, among other topics. Qualitative data on adverse experiences was collected online from participant households in 46 countries via and a separate survey | In response to the survey, more than 3 in 4 households reported an income loss since the beginning of the pandemic. Poorer households were more likely to suffer income losses (82%) than those not classified as poor (70%). Urban households were also disproportionately affected by income loss, with respondents from urban areas almost twice as likely to say they have lost their job (61%) compared with those in rural areas (33%). 70% of respondents who suffered economic losses reported that they had not received government |
|  |  |  |  |  | sent to populations of specific interest. | support. Nearly one third (32%) of households had a child, parent or caregiver who said that there had been physical or emotional violence in their home since the start of the pandemic. The number of households with a parent or caregiver respondent who said there was violence varied significantly across regions. |
| **Garstang, J., et al.** Effect of COVID-19 Lockdown on Child Protection Medical Assessments: A Retrospective Observational Study in Birmingham, UK. | Sep-20 | United Kingdom | Children under the age of 18 years | Quasi-experimental | Compared data for all child protection medical examinations (CPME) at a large, urban healthcare network for 18-week periods in 2018, 2019 and 2020 to compare changes in non-accidental injury (NAI) or neglect during the COVID-19 school shutdowns. Assessed child demographic data, referral details, and whether there were physical findings to support NAI or neglect. | The results indicate a 39.7% reduction in referrals for CPME from March to June 2020, compared to the same period in 2018 and a 37.3% reduction from 2019 to 2020. There were significantly fewer girls referred in 2020. The children referred for CPME in 2020 had similar social care histories to other years with the majority being previously known to social care and approximately half being open cases at the time of referral. |
| **Hansen, Alison L.** Child Maltreatment Reporting Statistics During the Covid-19 Pandemic: A cursory Analysis. | Aug-20 | USA | Children served by state protection agencies | Quasi-experimental | The number of total allegations of child maltreatment between the months of March and June—a span of time representative of the COVID-19 pandemic thus far—were analyzed in five different states in the years 2017, 2018, 2019, and 2020. | Utilizing publicly reported child maltreatment data from five states (Florida, New Jersey, Arizona, Nevada and Indiana), results reveal a disproportionate decline in child maltreatment reports during the COVID-19 pandemic (March to June 2020), compared to previous years from 2017 to 2019, across all five states. Indiana and Nevada experienced the greatest percentage change in reports of alleged child maltreatment. Nevada experienced a 26.6% decrease in reports between 2019 and 2020. Similarly, Indiana experienced a 24.3% decrease in reports of alleged child maltreatment in the same period. |
| **Jentsch, B., and B. Schnock.** Child Welfare in the Midst of the Coronavirus Pandemic-Emerging | Sep-20 | Germany | German- and English-language scientific | Review | A systematic search of German and English academic and gray literature using key word searches and the dates April 19, 2020 to August 27, 2020. Interim results of empirical | Results indicate that the number of reports on alleged child maltreatment had declined in March and April 2020. Out of 231 youth welfare agencies responding, 43% stated volume of reports had decreased in first four weeks following contact |
| Evidence from Germany. |  |  | articles, position papers and other gray literature |  | studies conducted in Germany were included. | restrictions. A more comprehensive survey of agencies carried out in May 2020 similarly reported that alleged child maltreatment reports had not increased and, in some cases, decreased. A similar development was noted concerning number of children being taken into care. |
| **Kovler, M. L., et al.**<br>Increased Proportion of Physical Child Abuse Injuries at a Level I Pediatric Trauma Center during the Covid-19 Pandemic. | Sep-20 | USA | Children under the age of 15 years | Quasi-experimental | Retrospective review of patients at a pediatric trauma centre to identify injuries caused by physical child abuse in the month following the statewide closure of childcare facilities in Maryland. The proportion of PCA patients treated during the Covid-19 era were compared to the corresponding period in the preceding two years. | Results indicate an increase in the proportion of traumatic injuries caused by physical child abuse in March and April 2020, compared to the same period in 2019 and 2018. Eight patients with physical child abuse injuries were treated during the Covid-19 period (13% of total trauma patients), compared to four in 2019 (4%) and three in 2018 (3%). The median age of patients in the Covid-19 period was 11.5 months. Most patients were black (75%) with public health insurance (75%). All injuries were caused by blunt trauma, resulting in scalp/face contusions (63%), skull fractures (50%), intracranial hemorrhage (38%), and long bone fractures (25%). |
| **Lawson, M., et al.**<br>Child Maltreatment during the COVID-19 Pandemic: Consequences of Parental Job Loss on Psychological and Physical Abuse towards Children. | Sep-20 | USA | *N* = 342 parents who are in a committed romantic relationship and who have a child age 4 to 10 years | Cross-sectional | Online survey data collected from mid-April to mid-May 2020, assessed experiences with COVID-19, the Parent-Child Conflict Tactics Scale, and the Family Crisis Oriented Personal Evaluation Scales. | Parental job loss was positively and significantly associated with psychological maltreatment and physical abuse towards children in the past week. Younger children were more likely to be physically abused and psychologically maltreated during the past week. Parents from families with more financial stability were less likely to physically abuse their children during the pandemic. Parents reporting more depressive symptoms were more likely to psychologically maltreat and physically abuse their children. Male children were more likely to be psychologically maltreated. |
| **Lee, S. J., and Ward, K. P.** Stress and | Mar-20 | USA | *N* = 288 parents with a | Cross-sectional | Online survey, conducted March 24-25, 2020 to investigate mental health and wellbeing, parenting, and | Parents are engaging in parent-child activities much more frequently since COVID-19. 81% of parents said that they praised or complimented their |
| Parenting During the Coronavirus Pandemic. |  |  | child age 0-12 that lives with them at least part of the time |  | economic situation during the pandemic. | child(ren) very often, and 88% of parents said they and their children had shown love to each other very often in the past 2 weeks. However, most parents (52%) said financial concerns and social isolation (50%) were getting in the way of their parenting. Most parents (61%) shouted, yelled or screamed at their children at least once in the past 2 weeks; 20% spanked or slapped their child at least once in the past 2 weeks. Parents reported they have yelled or screamed at their children more often (19%) or increased discipline (15%) since the Coronavirus pandemic |
| **Martins-Filho, P. R., et al.** Decrease in Child Abuse Notifications during COVID-19 Outbreak: A Reason for Worry or Celebration? | Sep-20 | Brazil | Children under the age of 18 years | Quasi-experimental | Compared data extracted from the official child maltreatment database (Coordination of Statistics and Criminal Analysis) for the first half of 2019 and 2020 in Sergipe state, Northeast Brazil. | The reported rate of child physical injury decreased in 2020 by 24.3% from 1 January to 30 June 2020, compared to the same time period in 2019. Reported rates were higher among girls (71.7%), children aged 12–17 years (69.8%) and Blacks/Browns (73.6%). These trends in child abuse notifications according to gender, age and race/ethnicity were not significantly different between 2020 and 2019. |
| **Øverlien, C. (2020).** The COVID-19 Pandemic and Its Impact on Children in Domestic Violence Refuges. | Aug-20 | Norway | N = 46<br>Staff at domestic violence refuges | Cross-sectional | An online survey was distributed to all domestic violence refuges in Norway from April 8-23, 2020, with the aim of obtaining an overview of the impact of the COVID-19 crisis and the virus control measures on victims of domestic violence and abuse. The survey contained 15 items with fixed answers, follow-up questions and an opportunity for the refuges to provide supplementary qualitative descriptions and examples. | 56% of respondents reported there had been a reduction in the number of requests from clients, many expressed concerns that it is 'too quiet out there.' Refuge staff reported being most concerned about children. Refuge staff were concerned that children exposed to violence who were living outside of the refuge would no longer receive the help that they need from professionals; and, moreover, that the violence they experience would remain undisclosed, as contact between adults outside of the family and the abused child is dramatically reduced. Closed schools and daycare centres are of particular concern as school represents normality and a zone free from the abusive parent. |
| **Rapoport, E., et al.** Reporting of Child Maltreatment During the SARS-CoV-2 Pandemic in New York City from March to May 2020. | Sep-20 | USA | Children under the age of 18 years | Quasi-experimental | Analyzed monthly data from New York City on the number of child maltreatment allegations using data from January 2015 to February 2020 to predict expected values for March, April, and May 2020. | The results show 29% fewer allegations of child maltreatment in March 2020 than forecasted. In April and May 2020, the number of reports was approximately half of what was expected. Significant decreases in child maltreatment reporting were also noted for all reporter subtypes including mandated reporters such as educational personnel, child care personnel and medical health personnel, and non-mandated reporters such as family members and neighbours. |
| **Ritz, D., et al.** The Hidden Impact of COVID-19 on Child Protection and Wellbeing. | Sep-20 | Multiple countries | $N = 17,565$<br>Parents and caregivers of a child under age 18 years | Cross-sectional | An online survey assessed implications of COVID-19 pandemic on Child Rights. Measured household-level characteristics, child rights, caregiver's well-being and family relationships, child education and well-being, among other topics. Sample included Save the Children program participants, specific population groups of interest to the program, and the general public. | Over 1 in 5 parents/caregivers (22%) surveyed reported an increase in their use of negative or violent parenting methods. 32% of households had a child and/or parent/caregiver reporting that violence had occurred in the home, including children and/or adults being verbally or physically abused. The vast majority of parents/caregivers (89%) reported an increase in negative feelings due to the COVID-19 pandemic. While 83% of the parents/caregivers reported an increase in negative feelings when schools were closed between 1 to 4 weeks, the reports of negative feelings were higher for the vast majority of adults (95%) when schools had been closed for 17 to 19 weeks. |
| **Sidpra, J., et al.** Rise in the Incidence of Abusive Head Trauma during the COVID-19 Pandemic. | Jun-20 | United Kingdom | Children under the age of 13 months | Quasi-experimental | Reviewed the incidence of suspected acute head trauma at an urban hospital between 23 March and 23 April 2020—the month that the United Kingdom entered a period of national self-isolation—and compared this with the incidence in the previous 3 years. | Review of the incidence of suspected abusive head trauma indicated a 1493% increase per month compared to the same period over the previous 3 years (10 cases in comparison with a mean of 0.67 cases per month). All families live in areas with a higher than average Index of Multiple Deprivation and 70% of parents had significant underlying vulnerabilities (e.g., criminal histories, mental health disorders, financial concerns). |
| **Tener, D., et al. “How Does COVID-19 Impact Intrafamilial Child Sexual Abuse? Comparison Analysis of Reports by Practitioners in Israel and the US.** | Oct-20 | USA and Israel | N = 50<br>Child welfare and legal professionals | Qualitative | An open-ended, online questionnaire distributed during May 2020 compare professional perspectives and experiences working with intrafamilial child sexual abuse with respect to the context of the COVID-19 pandemic within the United States and Israel. | Professionals in both countries were extremely concerned by the lack of opportunity to report the abuse. A surge in reports of intrafamilial child sexual abuse (IFCSA) was predicted after the COVID-19 period ends. Findings reveal mostly negative changes in the dynamics of IFCSA families during COVID-19, including financial, environmental, and emotional hardships, as well as some positive changes in relationships among family members. Risks of transferring to internet based or telephonic therapeutic interventions included technical issues, limited internet access among ultra-Orthodox families, lack of proper separate space for therapy, and difficulties engaging younger children. |
| **Toros, K., & Falch-Eriksen, A. A Child’s Right to Protection during the COVID-19 Crisis: An Exploratory Study of the Child Protective Services of Estonia.** | Oct-20 | Estonia | N = 81<br>Child protection workers | Qualitative | An online survey with three open-ended questions sent to child protection workers (CPW) in April and May 2020 collected case-worker narratives. Assessed the impact of COVID-19 on organisational design and, the workers’ ability to conduct a diagnosis of a child’s care context. | Results pointed towards challenges to facilitating children's protection during the pandemic. CPWs reported more difficulty identifying children and families in need; a decrease in referrals from the formal network (typically schools and kindergartens); perception of overload of the work by the networking partners; and a drop in provision of services for families in need, especially family therapy, psychology and child psychiatry. CPWs reported that child well-being and protection became a secondary consideration, and families' basic food needs and individual health protection from the virus became priorities. |
| **United Nations Children’s Fund. Protecting Children from Violence in the Time of COVID-19: Disruptions in Prevention and Response Services.** | Aug-20 | Multiple countries | N = 136 UNICEF country offices | Cross-sectional | Online survey collected information from May 1-August 14, 2020 from 136 UNICEF country offices on disruptions in service provision related to violence against children (VAC) as a result of the coronavirus. | South Asia has the highest proportion of countries reporting service disruptions, followed by Eastern Europe and Central Asia. Case management and home visits for children and women at risk of abuse are among the most commonly disrupted services. Around two thirds of countries with VAC-related service disruptions reported that at least one type of service had been severely affected. 70% of countries reported that mitigation measures had been put into place. |
| **Whaling, K. et al.**<br>Reduced Child Maltreatment Prevention Service Case Openings during COVID-19. | May-20<br>*Preprint* | USA | Children under the age of 18 years | Quasi-experimental | Analyzed data from New York City's Administration for Children's Services for January, February, and March for the years 2013-2020 to investigate the frequency of child maltreatment prevention service case openings. | An examination of trend lines demonstrated that new prevention case openings have consistently peaked in the month of March for 2013-2019. New prevention case service openings in March 2020 do not peak, as they do in the preceding seven years. Results show new child maltreatment prevention case openings in March 2020 were 61% lower than mean new case openings over March 2013–2019. |
| **World Vision.**<br>COVID-19 Rapid Impact Assessment Report. | May-20 | Bangladesh | N = 12,713 people, including adults (53.2%) and children (46.8%) | Mixed methods | Used mixed method approach to understand impact of COVID-19 on most vulnerable children, families and communities across Bangladesh. Data were quantitative and qualitative-household (HH) surveys, focus group discussion and KIIs with community leaders, children groups, government representatives and faith leaders. | Loss of incomes among households has triggered stress among parents and caregivers who are now engaging in desperate negative behaviours towards their children with about 35% applying physical punishment or psychological aggression. More than half of the respondent (58%) indicating that they were to partially able, with one-third (33.7%) indicating capability while 7.2% respondent expressed that they cannot handle the situation. |
| **World Vision.** Rapid Assessment of the Impact of COVID-19 on Child Wellbeing in Cambodia. | Jun-20 | Cambodia | N = 222 household heads (survey); 42 key informants (local leaders, etc.); 65 Agricultural Cooperative leaders (survey) | Mixed methods | The rapid assessment was conducted from May 11-14, 2020 to assess food insecurity; education; and livelihoods among vulnerable families. | 57% of parents and caregivers reported that parenting has become more complicated or challenging since the beginning of the COVID-19 outbreak and, as a result, only 32% of them reported that they could adequately manage their children's behaviour. A significant number of local authorities, child club leaders, faith leaders and members of Commune Committees for Women and Children (CCWCs) interviewed mentioned that they are concerned by domestic violence in the present context. However, the data from the household survey does not suggest a significant increase in the use of corporal punishments and additional research would be needed in this area to better assess the potential impact of the COVID-19 outbreak on domestic violence. |
| **World Vision.**<br>Unmasking the Impact of COVID-19 on Asia's Most Vulnerable Children. | Aug-20 | Bangladesh, Cambodia, India, Indonesia, Mongolia, Myanmar, Nepal, Philippines, and Sri Lanka | N = 19,065 parents/caregivers; and N = 14,669 children | Mixed methods | A household survey and child consultation were conducted in May 2020 via telephone and face to face to gain a better understanding on how the COVID-19 pandemic is impacting the lives of the most vulnerable children, their families and communities. | 27% of parents/caregivers indicated that they had resorted to physical punishment or emotional abuse in the last month (39% for urban parents) while 25% of the children confirmed that their parents or caregivers used physical punishment and/or psychological aggression in the past month. |

**Safety and Security: Child Injury**

Safety and Security: Child Injury
| Reference | Date Released | Setting | Population | Study Design | Data Source | Summary of Findings |
| --- | --- | --- | --- | --- | --- | --- |
| **Bram J. T, et al.**<br>Where Have All the Fractures Gone? The Epidemiology of Pediatric Fractures During | Sep-20 | USA | N = 1745 records of children under the age of 18 years | Quasi-experimental | Utilized hospital records from a level I pediatric trauma hospital from 15 March to 15 April 2020 to compare acute fractures during the COVID-19 pandemic with fractures | Results indicate that pediatric fracture volume decreased 2.5-fold during the COVID-19 pandemic. There was a significant decrease in the incidence of fractures (22.5 vs. 9.6 per day). The presenting age for all fractures decreased (7.5 vs. 9.4 years) because of decreased fracture burden among adolescents. There was also a decrease in the number of fractures requiring surgery (2.2 vs. 0.8 per day). During the pandemic, there was an |
| the COVID-19 Pandemic. |  |  |  |  | during a pre-pandemic period at the same institution. | increase in the proportion of injuries occurring at home (57.8% vs. 32.5%) or on bicycles (18.3% vs. 8.2%), but a decrease in those related to sports (7.2% vs. 26.0%) or playgrounds (5.2% vs. 9.0%). |
| **Chaiyachati, B. H., et al.** Trends in Pediatric Emergency Department Utilization after Institution of COVID-19 Mandatory Social Distancing. | Jul-20 | USA | N = 231 records of children under age 21 years | Quasi-experimental | Compared emergency department (ED) visits to a tertiary urban children's hospital from January to April 2020 with the same date range during the 3 prior years (2017-2019). | Pediatric emergency healthcare use from January to April 2020 compared with the prior 3 years indicates a rapid decrease in emergency department visits (95 vs 286). Trauma, fever, and abdominal pain had higher proportions of high-acuity presentations in 2020. The average number of ingestions was higher in 2020 than in the comparison period (31 vs 25). The proportion of patients categorized as high acuity was higher in 2020 (59.4% vs 49.6%). Similarly, the proportions of patients admitted both overall (22.4% vs 18.5%) and to the intensive care unit (2.8% vs 1.7%) were higher in 2020. |
| **Hamill J. K. and Sawyer M C.** Reduction of childhood trauma during the COVID-19 Level 4 lockdown in New Zealand. | Jun-20 | New Zealand | Children under the age of 18 years | Quasi-experimental | Compared patient data from New Zealand's only designated paediatric major trauma centre from period of lockdown from 26 March to 27 April 2020, to the same date range from the years 2016 to 2019 | Results show total trauma admissions during the lockdown were at a 5-year low. There were 57 trauma admissions (four major trauma) during the lockdown compared to an average of 85 (six major trauma) during the same time period in previous years. No motor vehicle crash occupants were admitted. Nineteen percent of children were injured on bicycles compared to an average of 3.6% in previous years. |
| **Li, H., et al.** Changes in Children's Healthcare Visits During Coronavirus Disease-2019 Pandemic in Hangzhou, China. | May-20 | China | Children under the age of 18 years | Quasi-experimental | Compared data from outpatient healthcare visits between January 1, 2020, and March 31, 2020, as well as historical data during the same period of 2019 for a tertiary Children's Hospital in a large Chinese City. | Coincident with COVID-19 restrictions, the daily number of pediatric visits after January 25, 2020, decreased to about only one-quarter compared with the historical data in 2019. The number of injuries also rose significantly, especially indoor injuries such as finger injuries, foreign body entering eye or natural orifice, tibial fracture, eyelid laceration, and supracondylar fracture of humerus. Some cases of epistaxis also were due to injuries. After January 25, 2020, there were 19 electrical injuries seen, compared with only 6 in the first 3 months in 2019. |
| **Nabian M. H., et al.** Epidemiological Pattern of Pediatric Trauma in COVID-19 Outbreak: Data from a Tertiary Trauma Center in Iran. | Sep-20 | Iran | Children under the age of 18 years | Quasi-experimental | Compared patient data on pediatric fractures from a tertiary trauma center from March 1-April 15, 2020 with mean data from the past 2 years. | 117 of 288 trauma patients during the 2020 period (40.62%) had a fractured bone (145 fractures). Patients were mostly boys, with a mean age of 9.87 years. The three most common fracture types in children involved arm fractures. Compared to non-COVID era, the number of pediatric trauma admissions dropped from 589 to 288. No significant change happened in the mean age, male/female ratio and percentage of motor vehicle accidents. Proportion of proximal humeral, proximal forearm, carpal, and hand fractures declined. The number of open fractures significantly dropped (from 12 to 2). |
| **Raitio A, et al.** Reduced Number of Pediatric Orthopedic Trauma Requiring Operative Treatment during COVID-19 Restrictions: A Nationwide Cohort Study. | Oct-20 | Finland | Children under the age of 16 years | Quasi-experimental | Assessed pediatric emergency operations related to fractures, appendicitis and scrotal exploration for all five tertiary centers in Finland for the periods from March 1-May 31 for 2017-2020. | Results show a significant decrease (31%) in trauma operations compared to the same period during the previous three years. Daycare, school, and organized sports-related injuries decreased. The frequencies for scrotal exploration and surgery for acute appendicitis were similar during the pre-pandemic and pandemic era. There was no change in the age or sex of the children undergoing emergency surgery in four university hospitals; in one hospital, the age of the fracture patients was significantly lower during the pandemic months (median 12.2 versus 10.4 years). |
| **Sugand K, et al.** Impact of the COVID-19 Pandemic on Paediatric Orthopaedic Trauma Workload in Central London: A Multi-centre Longitudinal Observational Study over the “Golden Weeks” | Aug-20 | UK | Children under the age of 18 years | Quasi-experimental | Compared patient data from a large acute paediatric hospital service in urban UK during the first lockdown from 17 March to 28 April 2020 with the same period in 2019, including both acute paediatric orthopaedic trauma referrals and operative caseload. | There was a significant reduction in the odds of sporting-related mechanism of injuries (by 57%) in 2020 compared with 2019. There were only 97 referrals following the introduction of social distancing measures in 2020 compared with 302 in 2019. This represents a 68% reduction in paediatric injuries. The general demographic of those presenting with injuries changed between the 2 periods, with a significantly younger median age in 2020 and more girls. The pattern of injury also remained generally unchanged with upper limb injuries being the most common at 67% and 69% respectively in 2019 and 2020. In 2020 there were 3% more safeguarding referrals for non-accidental injuries (not a statistically significant difference compared to |
|  |  |  |  |  |  | 2019). There has also not been a large increase in delayed presentation of substantial injuries. |
| **Turgut A, et al.**<br>Effect of COVID-19 Pandemic on the Fracture Demographics: Data from a Tertiary Care Hospital in Turkey. | Jul-20 | Turkey | Children under the age of 17 years | Quasi-experimental | Assessed records of patients admitted to a tertiary care hospital with a new fracture during pandemic period of March 16 to May 22, 2020, compared to the same date range in 2018 and 2019. Compared pediatric ($\leq 16$ years) and non-pediatric ($> 16$ years) | A total of 1794, 1747, and 670 fractures were observed in 2018, 2019, and 2020, respectively. Mean age of the patients in pediatric group was found to have decreased significantly in the pandemic period. The most common fracture sites in the pediatric population were the distal forearm and distal arm. The proportional increase in femoral and tibial shaft fractures in the pediatric group was found to be statistically significantly. In the pediatric group 6.8%, 7.7%, and 14.6% of the fractures were treated surgically in 2018, 2019, and 2020, respectively. |
| **Verdoni F., et al.**<br>Effect of the COVID-19 Outbreak on Pediatric Patients' Admissions to the Emergency Department in an Italian Orthopedic Trauma Hub. | Sep-20 | Italy | Children under the age of 17 years | Quasi-experimental | The records of pediatric patients admitted to emergency room of an orthopedic trauma center during the pandemic period from 12 March to 5 May 2020 was compared to the same period in 2019. | Results show a reduction of 87% in admissions. A statistically significant difference between the two groups was found in the mean age of patients, that decreased from 11.4 prior to the pandemic to 8.6 years during the pandemic. a trend towards more severe codes was observed in the pandemic group and the diagnosis of fracture was significantly less frequent in non-pandemic group and injuries at home were more frequent (34.8% compared to 6.8% in the non-pandemic group). |
| **Williams F. N., et al.** COVID-19 related Admissions to a Regional Burn Center: The impact of Shelter-in-place Mandate. | Oct-20 | USA | School-aged children from age 6 to 17 years | Quasi-experimental | Compared all burn center admissions from between 10 March and 22 May 2020 to a month-matched historical cohort from 2019 and summer-time admissions for the last five years. | Compared to the 2019 cohort, there was a 28% increase in admissions of school aged children in 2020 during the shelter-in-place order time frame. Comparing the percentage of school age admissions from 10 March to 22 May 2020 (11%) to the percentage of school age admissions during the summer breaks from 2015 to 2019, the percentages were essentially equivalent (11%, 11%, 12%, 10%, 10%, respectively). None of these comparisons were statistically significant. There were also no statistically significant differences in race, sex, or type of insurance. For school-age children, there was a 36% increase in those with free or low cost federal and state health insurance in 2020 compared to 2019, though this did not reach statistical significance. |

## REFERENCES

1. Cluver L, Lachman JM, Sherr L, et al. Parenting in a time of COVID-19. Published Online First: 2020. doi:10.1016/S0140-6736(20)30736-4

2. Yoshikawa H, Wuermli AJ, Britto PR, et al. Effects of the Global Coronavirus Disease-2019 Pandemic on Early Childhood Development: Short- and Long-Term Risks and Mitigating Program and Policy Actions. J Pediatr 2020;223:188–93. doi:10.1016/j.jpeds.2020.05.020

3. UNICEF, World Bank, World Health Organization. Nurturing care for early childhood development a framework for helping children survive and thrive to transform health and human potential. Geneva: World Health Organization 2018. https://www.who.int/maternal_child_adolescent/documents/nurturing-care-early-childhood-development/en (accessed 8 Jul 2020).

4. Chow MYK, Morrow AM, Booy R, et al. Impact of children’s influenza-like illnesses on parental quality of life: a qualitative study. J Paediatr Child Health 2013;49:664–70. doi:10.1111/jpc.12261

5. Effler PV, Carcione D, Giele C, et al. Household responses to pandemic (H1N1) 2009-related school closures, Perth, Western Australia. Emerg Infect Dis 2010;16:205–11. doi:10.3201/eid1602.091372

6. Sprang G, Silman M. Posttraumatic stress disorder in parents and youth after health-related disasters. Disaster Med Public Health Prep 2013;7:105–10. doi:10.1017/dmp.2013.22

7. Green E, Chase RM, Zayzay J, et al. The impact of the 2014 Ebola virus disease outbreak in Liberia on parent preferences for harsh discipline practices: a quasi-experimental, pre-post design. Glob Ment Health Camb Engl 2018;5:e1. doi:10.1017/gmh.2017.24

8. Kodish SR, Bio F, Oemcke R, et al. A qualitative study to understand how Ebola Virus Disease affected nutrition in Sierra Leone—A food value-chain framework for improving future response strategies. PLoS Negl Trop Dis 2019;13:e0007645. doi:10.1371/journal.pntd.0007645

9. Plan International. Ebola: Beyond the Health Emergency. Plan International 2015. https://plan-international.org/publications/ebola-beyond-health%C2%A0emergency (accessed 4 Dec 2020).

10. Adibellİ D, SÜmen A. The Effect of the Coronavirus (Covid-19) Pandemic on Health-Related Quality of Life in Children. Child Youth Serv Rev 2020;119:105595. doi:10.1016/j.childyouth.2020.105595

11. Asbury K, Fox L, Deniz E, et al. How is COVID-19 Affecting the Mental Health of Children with Special Educational Needs and Disabilities and Their Families? J Autism Dev Disord 2020;:1–9. doi:10.1007/s10803-020-04577-2

12. Asia-Pacific Regional Network for Early Childhood. Perspectives on the impact of COVID-19 on young children and early childhood development in the Asia-Pacific Region. 2020.

13. Brown SM, Doom JR, Lechuga-Peña S, et al. Stress and parenting during the global COVID-19 pandemic. Child Abuse Negl 2020;:104699. doi:10.1016/j.chiabu.2020.104699

14. Cameron EE, Joyce KM, Delaquis CP, et al. Maternal psychological distress & mental health service use during the COVID-19 pandemic. J Affect Disord 2020;276:765–74. doi:10.1016/j.jad.2020.07.081

15. Ceulemans M, Hompes T, Foulon V. Mental health status of pregnant and breastfeeding women during the COVID-19 pandemic: A call for action. Int J Gynaecol Obstet Published Online First: 3 July 2020. doi:10.1002/ijgo.13295

16. Chung G, Lanier P, Wong PYJ. Mediating Effects of Parental Stress on Harsh Parenting and Parent-Child Relationship during Coronavirus (COVID-19) Pandemic in Singapore. J Fam Violence 2020;:1–12. doi:10.1007/s10896-020-00200-1

17. Davenport MH, Meyer S, Meah VL, et al. Moms Are Not OK: COVID-19 and Maternal Mental Health. Front Glob Womens Health 2020;1:1. doi:10.3389/fgwh.2020.00001

18. Dhiman S, Sahu PK, Reed WR, et al. Impact of COVID-19 outbreak on mental health and perceived strain among caregivers tending children with special needs. Res Dev Disabil 2020;107:103790. doi:10.1016/j.ridd.2020.103790

19. Di Giorgio E, Di Riso D, Mioni G, et al. The interplay between mothers’ and children behavioral and psychological factors during COVID-19: an Italian study. Eur Child Adolesc Psychiatry Published Online First: 31 August 2020. doi:10.1007/s00787-020-01631-3

20. Dib S, Rougeaux E, Vázquez-Vázquez A, et al. Maternal mental health and coping during the COVID-19 lockdown in the UK: Data from the COVID-19 New Mum Study. Int J Gynaecol Obstet Published Online First: 26 September 2020. doi:10.1002/ijgo.13397

21. Dodd H, Westbrook J, Lawrence P. Co-SPYCE Report One: Findings from 1728 parents/carers of 2-4 year olds on stress, child activities, child worries and need for support. 2020;:10.

22. Evans S, Mikocka-Walus A, Klas A, et al. From “It Has Stopped Our Lives” to “Spending More Time Together Has Strengthened Bonds”: The Varied Experiences of Australian Families During COVID-19. Front Psychol 2020;11. doi:10.3389/fpsyg.2020.588667

23. Farewell CV, Jewell J, Walls J, et al. A Mixed-Methods Pilot Study of Perinatal Risk and Resilience During COVID-19. J Prim Care Community Health 2020;11:2150132720944074. doi:10.1177/2150132720944074

24. Gassman-Pines A, Ananat EO, Fitz-Henley J. COVID-19 and Parent-Child Psychological Well-being. Pediatrics 2020;146:e2020007294. doi:10.1542/peds.2020-007294

25. Hamadani JD, Hasan MI, Baldi AJ, et al. Immediate impact of stay-at-home orders to control COVID-19 transmission on socioeconomic conditions, food insecurity, mental health, and intimate partner violence in Bangladeshi women and their families: an interrupted time series. Lancet Glob Health 2020;8:e1380–9. doi:10.1016/s2214-109x(20)30366-1

26. Hiraoka D, Tomoda A. Relationship between parenting stress and school closures due to the COVID-19 pandemic. Psychiatry Clin Neurosci Published Online First: 17 June 2020. doi:10.1111/pcn.13088

27. Kalil A, Mayer S, Shah R. Impact of the COVID-19 Crisis on Family Dynamics in Economically Vulnerable Households. Harris School of Public Policy Studies, University of Chicago 2020.

28. Lee S, Ward K. Stress and Parenting During the Coronavirus Pandemic. University of Michigan 2020.

29. Limbers CA, McCollum C, Greenwood E. Physical activity moderates the association between parenting stress and quality of life in working mothers during the COVID-19 pandemic. Ment Health Phys Act 2020;19:100358. doi:10.1016/j.mhpa.2020.100358

30. Marchetti D, Fontanesi L, Mazza C, et al. Parenting-Related Exhaustion During the Italian COVID-19 Lockdown. J Pediatr Psychol Published Online First: 17 October 2020. doi:10.1093/jpepsy/jsaa093

31. Mazza C, Ricci E, Marchetti D, et al. How Personality Relates to Distress in Parents during the Covid-19 Lockdown: The Mediating Role of Child’s Emotional and Behavioral Difficulties and the Moderating Effect of Living with Other People. Int J Env Res Public Health 2020;17. doi:10.3390/ijerph17176236

32. McRae CS, Henderson AM, Low RS, et al. Parents’ Distress and Poor Parenting during COVID-19: The Buffering Effects of Partner Support and Cooperative Coparenting. Published Online First: 2020.https://psyarxiv.com/nxdsk/

33. Nolvi S, Karukivi M, Korja R, et al. Parental depressive and anxiety symptoms as a response to the COVID-19 pandemic: a birth cohort follow-up study. PsyArXiv 2020. doi:10.31234/osf.io/8htb4

34. Oskovi-Kaplan ZA, Buyuk GN, Ozgu-Erdinc AS, et al. The Effect of COVID-19 Pandemic and Social Restrictions on Depression Rates and Maternal Attachment in Immediate Postpartum Women: a Preliminary Study. Psychiatr Q 2020;:1–8. doi:10.1007/s11126-020-09843-1

35. Patrick SW, Henkhaus LE, Zickafoose JS, et al. Well-being of Parents and Children During the COVID-19 Pandemic: A National Survey. Pediatrics 2020;146. doi:10.1542/peds.2020-016824

36. Peltz JS, Daks JS, Rogge RD. Mediators of the association between COVID-19-related stressors and parents’ psychological flexibility and inflexibility: The roles of perceived sleep quality and energy. J Context Behav Sci 2020;17:168–76. doi:10.1016/j.jcbs.2020.07.001

37. Romero E, López-Romero L, Domínguez-Álvarez B, et al. Testing the Effects of COVID-19 Confinement in Spanish Children: The Role of Parents’ Distress, Emotional Problems and Specific Parenting. Int J Environ Res Public Health 2020;17. doi:10.3390/ijerph17196975

38. Roos LE, Salisbury M, Penner-Goeke L, et al. Supporting Families to Protect Child Health: Parenting Quality and Household Needs During the COVID-19 Pandemic. Published Online First: 2020. doi:10.31234/osf.io/u5xzw

39. Russell BS, Hutchison M, Tambling R, et al. Initial Challenges of Caregiving During COVID-19: Caregiver Burden, Mental Health, and the Parent-Child Relationship. Child Psychiatry Hum Dev 2020;51:671–82. doi:10.1007/s10578-020-01037-x

40. Spinelli M, Lionetti F, Pastore M, et al. Parents’ Stress and Children’s Psychological Problems in Families Facing the COVID-19 Outbreak in Italy. Front Psychol 2020;11:1713. doi:10.3389/fpsyg.2020.01713

41. Spinelli M, Lionetti F, Setti A, et al. Parenting Stress During the COVID-19 Outbreak: Socioeconomic and Environmental Risk Factors and Implications for Children Emotion Regulation. Fam Process Published Online First: 28 September 2020. doi:10.1111/famp.12601

42. Westrupp E, Bennett C, Berkowitz TS, et al. Child, parent, and family mental health and functioning in Australia during COVID-19: Comparison to pre-pandemic data. PsyArXiv 2020. doi:10.31234/osf.io/ydrm9

43. Willner P, Rose J, Kroese BS, et al. Effect of the COVID-19 pandemic on the mental health of carers of people with intellectual disabilities. J Appl Res Intellect Disabil 2020;33:1523– 33. doi:https://doi.org/10.1111/jar.12811

44. Xu Y, Wu Q, Levkoff SE, et al. Material hardship and parenting stress among grandparent kinship providers during the COVID-19 pandemic: The mediating role of grandparents’ mental health. Child Abuse Negl 2020;:104700. doi:10.1016/j.chiabu.2020.104700

45. Zanardo V, Manghina V, Giliberti L, et al. Psychological impact of COVID-19 quarantine measures in northeastern Italy on mothers in the immediate postpartum period. Int J Gynaecol Obstet 2020;150:184–8. doi:10.1002/ijgo.13249

46. Brandstetter S, Poulain T, Vogel M, et al. The impact of the COVID-19 pandemic on families in Germany. medRxiv 2020;:2020.10.05.20206805. doi:10.1101/2020.10.05.20206805

47. Loperfido L, Burgess M. The Hidden Impact of COVID-10 on Child Povery. London: Save the Children International 2020. https://resourcecentre.savethechildren.net/node/18174/pdf/the_hidden_impact_of_covid-19_on_child_poverty.pdf

48. Ritz D, O’Hare G, Burgess M. The Hidden Impact of COVID-19 on Child Protection and Wellbeing. London: Save the Children International 2020. https://resourcecentre.savethechildren.net/node/18174/pdf/the_hidden_impact_of_covid-19_on_child_protection_and_wellbeing.pdf

49. World Vision. COVID-19 rapid impact assessment report. 2020. https://www.wvi.org/sites/default/files/2020-07/Rapid%20Impact%20Assesment%20Report_Full%20Report.pdf

50. Chung G, Chan XW, Lanier P, et al. Associations Between Work-Family Balance, Parenting Stress, and Marital Conflicts During COVID-19 Pandemic in Singapore. Open Science Framework 2020. doi:10.31219/osf.io/nz9s8

51. Joyce KM, Cameron E, Slymka J, et al. Changes in Maternal Substance Use During the COVID-19 Pandemic. Published Online First: 2020.https://psyarxiv.com/htny8/

52. Abdelrahman M, Al-Adwan D, Hassan Y. Impact of social distancing on the mental health of parents and children in Qatar. Published Online First: 2020. doi:10.31219/osf.io/qpu6d

53. Chu K, Schwartz C, Towner E, et al. Parenting Under Pressure: A Mixed-Methods Investigation of the Impact of COVID-19 on Family Life. PsyArXiv 2020. doi:10.31234/osf.io/zm39b

54. Günther-Bel C, Vilaregut A, Carratala E, et al. A Mixed-method Study of Individual, Couple, and Parental Functioning During the State-regulated COVID-19 Lockdown in Spain. Fam Process 2020;59:1060–79. doi:10.1111/famp.12585

55. Brown A, Shenker N. Experiences of breastfeeding during COVID-19: Lessons for future practical and emotional support. Matern Child Nutr 2020;:e13088. doi:10.1111/mcn.13088

56. Das R. COVID-19, perinatal mental health and the digital pivot. 2020;:32.

57. Saunders B, Hogg S. Babies in Lockdown: Listening to parent to build back better. 2020. https://parentinfantfoundation.org.uk/our-work/campaigning/babies-in-lockdown/#fullreport

58. Spatz DL, Froh EB. Birth and Breastfeeding in the Hospital Setting during the COVID-19 Pandemic. MCN Am J Matern Child Nurs Published Online First: 9 October 2020. doi:10.1097/nmc.0000000000000672

59. Vazquez-Vazquez A, Dib S, Rougeaux E, et al. The impact of the Covid-19 lockdown on the experiences and feeding practices of new mothers in the UK: Preliminary data from the COVID-19 New Mum Study. Appetite 2020;:104985. doi:10.1016/j.appet.2020.104985

60. Andrew A, Cattan S, Dias MC, et al. How are mothers and fathers balancing work and family under lockdown?”. Inst Fisc Stud 2020.

61. Carlson DL, Petts R, Pepin J. Changes in Parents’ Domestic Labor During the COVID-19 Pandemic. SocArXiv 2020. doi:10.31235/osf.io/jy8fn

62. Craig L, Churchill B. Dual-earner parent couples’ work and care during COVID-19. Gend Work Organ 2020;:1–14. doi:https://doi.org/10.1111/gwao.12497

63. Del Boca D, Oggero N, Profeta P, et al. Women’s and men’s work, housework and childcare, before and during COVID-19. Rev Econ Househ Published Online First: 6 September 2020. doi:10.1007/s11150-020-09502-1

64. Farré L, Fawaz Y, González L, et al. How the covid-19 lockdown affected gender inequality in paid and unpaid work in spain. Published Online First: 2020.https://www.iza.org/publications/dp/13434/how-the-covid-19-lockdown-affected-gender-inequality-in-paid-and-unpaid-work-in-spain

65. Lyttelton T, Zang E, Musick K. Gender Differences in Telecommuting and Implications for Inequality at Home and Work. 2020.

66. Mangiavacchi L, Piccoli L, Pieroni L. Fathers Matter: Intra-Household Responsibilities and Children’s Wellbeing during the COVID-19 Lockdown in Italy. IZA Discussion Paper 2020.

67. Zamarro G, Prados M. Gender Differences in Couples’ Division of Childcare, Work and Mental Health During COVID-19. 2020.

68. Hjálmsdóttir A, Bjarnadóttir VS. ‘I have turned into a foreman here at home.’ Families and work-life balance in times of Covid-19 in a gender equality paradise. Gend Work Organ Published Online First: 19 September 2020. doi:10.1111/gwao.12552

69. Heggeness M. Estimating the Immediate Impact of the COVID-19 Shock on Parental Attachment to the Labor Market and the Double Bind of Mothers. Rev Econ Househ 2020.

70. Manzo LKC, Minello A. Mothers, childcare duties, and remote working under COVID-19 lockdown in Italy: Cultivating communities of care. Dialogues Hum Geogr 2020;10:120–3. doi:10.1177/2043820620934268

71. Qian Y, Fuller S. COVID-19 and the Gender Employment Gap among Parents of Young Children. Can Public Policy 2020;46:S89–101. doi:10.3138/cpp.2020-077

72. Barnett S, Jung K, Nores M. Young Children’s Home Learning and Preschool Participation Experiences During the Pandemic: NIEER 2020 Preschool Learning Activities Survey: Technical Report and Selected Findings. National Institute for Early Education Research 2020. http://nieer.org/wp-content/uploads/2020/08/NIEER-Tech-Rpt-July-2020-Young-Children%E2%80%99s-Home-Learning-and-Preschool-Participation-Experiences-During-the-Pandemic-AUG-2020-1.pdf

73. Kim J, Araya M, Ejigu C, et al. The Implications of COVID-19 on early learning continuity in Ethiopia: Perspectives of parents and caregivers. REAL Centre, University of Cambridge 2020.

74. Dong C, Cao S, Li H. Young children’s online learning during COVID-19 pandemic: Chinese parents’ beliefs and attitudes. Child Youth Serv Rev 2020;118:105440. doi:10.1016/j.childyouth.2020.105440

75. Briggs DC. COVID-19: The Effect of Lockdown on Children’s Remote Learning Experience – Parents’ Perspective. J Educ Soc Behav Sci 2020;:42–52. doi:10.9734/jesbs/2020/v33i930257

76. Grover S, Goyal SK, Mehra A, et al. A Survey of Parents of Children Attending the Online Classes During the Ongoing COVID-19 Pandemic. Indian J Pediatr Published Online First: 10 October 2020. doi:10.1007/s12098-020-03523-5

77. Dayal HC, Tiko L. When are we going to have the real school? A case study of early childhood education and care teachers’ experiences surrounding education during the COVID-19 pandemic. Australas J Early Child 2020;45:336–47. doi:10.1177/1836939120966085

78. World Vision. Rapid assessment of the impact of COVID-19 on child wellbeing in Cambodia. 2020. https://www.wvi.org/sites/default/files/2020-06/Rapid%20assessment%20Covid%2019%20report-02June20.pdf

79. Lee SJ, Ward KP, Chang OD, et al. Parenting Activities and the Transition to Home-based Education During the COVID-19 Pandemic. Child Youth Serv Rev 2020;:105585. doi:10.1016/j.childyouth.2020.105585

80. Drouin M, McDaniel BT, Pater J, et al. How Parents and Their Children Used Social Media and Technology at the Beginning of the COVID-19 Pandemic and Associations with Anxiety. Cyberpsychol Behav Soc Netw Published Online First: 27 July 2020. doi:10.1089/cyber.2020.0284

81. Carroll N, Sadowski A, Laila A, et al. The Impact of COVID-19 on Health Behavior, Stress, Financial and Food Security among Middle to High Income Canadian Families with Young Children. Nutrients 2020;12. doi:10.3390/nu12082352

82. Dunton GF, Do B, Wang SD. Early effects of the COVID-19 pandemic on physical activity and sedentary behavior in children living in the U.S. BMC Public Health 2020;20:1351. doi:10.1186/s12889-020-09429-3

83. Moore SA, Faulkner G, Rhodes RE, et al. Impact of the COVID-19 virus outbreak on movement and play behaviours of Canadian children and youth: a national survey. Int J Behav Nutr Phys Act 2020;17:85. doi:10.1186/s12966-020-00987-8

84. de Lannoy L, Rhodes RE, Moore SA, et al. Regional differences in access to the outdoors and outdoor play of Canadian children and youth during the COVID-19 outbreak. Can J Public Health Published Online First: 14 October 2020. doi:10.17269/s41997-020-00412-4

85. Xiang M, Zhang Z, Kuwahara K. Impact of COVID-19 pandemic on children and adolescents’ lifestyle behavior larger than expected. Prog Cardiovasc Dis 2020;63:531–2. doi:10.1016/j.pcad.2020.04.013

86. Mitra R, Moore SA, Gillespie M, et al. Healthy movement behaviours in children and youth during the COVID-19 pandemic: Exploring the role of the neighbourhood environment. Health Place 2020;65:102418. doi:10.1016/j.healthplace.2020.102418

87. Idoiaga Mondragon N, Berasategi Sancho N, Dosil Santamaria M, et al. Struggling to breathe: a qualitative study of children’s wellbeing during lockdown in Spain. Psychol Health 2020;:1–16. doi:10.1080/08870446.2020.1804570

88. Guerrero MD, Vanderloo LM, Rhodes RE, et al. Canadian children’s and youth’s adherence to the 24-h movement guidelines during the COVID-19 pandemic: A decision tree analysis. J Sport Health Sci 2020;9:313–21. doi:10.1016/j.jshs.2020.06.005

89. Barboza GE, Schiamberg LB, Pachl L. A spatiotemporal analysis of the impact of COVID-19 on child abuse and neglect in the city of Los Angeles, California. Child Abuse Negl 2020;:104740. doi:10.1016/j.chiabu.2020.104740

90. Bullinger LR, Carr JB, Packham A. COVID-19 and Crime: Effects of Stay-at-Home Orders on Domestic Violence. UNICEF Innocenti library: 2020. https://apackham.github.io/mywebsite/COVID_crime_webversion.pdf

91. Bullinger L, Boy A, Feely M, et al. COVID-19 and Alleged Child Maltreatment. Rochester, NY: Social Science Research Network 2020. doi:10.2139/ssrn.3702704

92. Martins-Filho PR, Damascena NP, Lage RC, et al. Decrease in child abuse notifications during COVID-19 outbreak: A reason for worry or celebration? J Paediatr Child Health Published Online First: 4 October 2020. doi:10.1111/jpc.15213

93. Baron EJ, Goldstein EG, Wallace CT. Suffering in Silence: How COVID-19 School Closures Inhibit the Reporting of Child Maltreatment. Published Online First: 2020. doi:10.2139/ssrn.3601399

94. Garstang J, Debelle G, Anand I, et al. Effect of COVID-19 lockdown on child protection medical assessments: a retrospective observational study in Birmingham, UK. BMJ Open 2020;10:e042867. doi:10.1136/bmjopen-2020-042867

95. Whaling K, Sarkissian AD, Larez NA, et al. Reduced child maltreatment prevention service case openings during COVID-19. In Review 2020. doi:10.21203/rs.3.rs-30930/v1

96. Rapoport E, Reisert H, Schoeman E, et al. Reporting of Child Maltreatment During the SARS-CoV-2 Pandemic in New York City from March to May 2020. Child Abuse Negl 2020;:104719. doi:10.1016/j.chiabu.2020.104719

97. Cabrera-Hernández F, Padilla-Romo M. Hidden Violence: How COVID-19 School Closures reduced the Reporting of Child Maltreatment. University of Tennessee, Department of Economics: 2020. https://ideas.repec.org/p/ten/wpaper/2020-02.html

98. Hansen AL. Child Maltreatment Reporting Statistics During the Covid-19 Pandemic: A Cursory Analysis. UNICEF Innocenti Library: Center for Health Law Policy and Bioethics Scholarship 2020. https://digital.sandiego.edu/cgi/viewcontent.cgi?article=1080&context=law_chlb_research_scholarship

99. Toros K, Falch-Eriksen A. A child’s right to protection during the COVID-19 crisis: An exploratory study of the Child Protective Services of Estonia. Child Youth Serv Rev 2020;119:105568. doi:10.1016/j.childyouth.2020.105568

100. Jentsch B, Schnock B. Child welfare in the midst of the coronavirus pandemic-Emerging evidence from Germany. Child Abuse Negl 2020;:104716. doi:10.1016/j.chiabu.2020.104716

101. Bhopal S, Buckland A, McCrone R, et al. Who has been missed? Dramatic decrease in numbers of children seen for child protection assessments during the pandemic. Arch Child Published Online First: 18 June 2020. doi:10.1136/archdischild-2020-319783

102. Øverlien C. The COVID-19 Pandemic and Its Impact on Children in Domestic Violence Refuges. Child Abuse Rev Published Online First: 18 August 2020. doi:10.1002/car.2650

103. Tener D, Marmor A, Katz C, et al. How does COVID-19 impact intrafamilial child sexual abuse? Comparison analysis of reports by practitioners in Israel and the US. Child Abuse Negl 2020;:104779. doi:10.1016/j.chiabu.2020.104779

104. Kovler ML, Ziegfeld S, Ryan LM, et al. Increased proportion of physical child abuse injuries at a level I pediatric trauma center during the Covid-19 pandemic. Child Abuse Negl 2020;:104756. doi:10.1016/j.chiabu.2020.104756

105. Sidpra J, Abomeli D, Hameed B, et al. Rise in the incidence of abusive head trauma during the COVID-19 pandemic. Arch Dis Child 2020;:archdischild-2020-319872. doi:10.1136/archdischild-2020-319872

106. Lawson M, Piel MH, Simon M. Child Maltreatment during the COVID-19 Pandemic: Consequences of Parental Job Loss on Psychological and Physical Abuse Towards Children. Child Abuse Negl 2020;:104709. doi:10.1016/j.chiabu.2020.104709

107. Bram JT, Johnson MA, Magee LC, et al. Where Have All the Fractures Gone? The Epidemiology of Pediatric Fractures During the COVID-19 Pandemic. J Pediatr Orthop 2020;40:373–9. doi:10.1097/BPO.0000000000001600

108. Chaiyachati B, Agawu A, Zorc J, et al. Trends in Pediatric Emergency Department Utilization after Institution of COVID-19 Mandatory Social Distancing. J Pediatr 2020;0.

109. Hamill JK, Sawyer MC. Reduction of childhood trauma during the COVID-19 Level 4 lockdown in New Zealand. Anz J Surg Published Online First: 26 June 2020. doi:10.1111/ans.16108

110. Nabian MH, Vosoughi F, Najafi F, et al. Epidemiological pattern of pediatric trauma in COVID-19 outbreak: Data from a tertiary trauma center in Iran. Injury Published Online First: 16 September 2020. doi:10.1016/j.injury.2020.09.015

111. Sugand K, Park C, Morgan C, et al. Impact of the COVID-19 pandemic on paediatric orthopaedic trauma workload in central London: a multi-centre longitudinal observational study over the “golden weeks”. Acta Orthop 2020;0:1–6. doi:10.1080/17453674.2020.1807092

112. Raitio A, Ahonen M, Jääskelä M, et al. Reduced Number of Pediatric Orthopedic Trauma Requiring Operative Treatment during COVID-19 Restrictions: A Nationwide Cohort Study. Scand J Surg 2020;:1457496920968014. doi:10.1177/1457496920968014

113. Turgut A, Arlı H, Altundağ Ü, et al. Effect of COVID-19 pandemic on the fracture demographics: Data from a tertiary care hospital in Turkey. Acta Orthop Traumatol Turc 2020;54:355–63. doi:10.5152/j.aott.2020.20209

114. Li H, Yu G, Duan H, et al. Changes in Children’s Healthcare Visits During Coronavirus Disease-2019 Pandemic in Hangzhou, China. J Pediatr 2020;224:146–9. doi:10.1016/j.jpeds.2020.05.013

115. Verdoni F, Ricci M, Di grigoli C, et al. Effect of the COVID-19 Outbreak on Pediatric Patients’ Admissions to the Emergency Department in an Italian Orthopedic Trauma Hub. Published Online First: 18 September 2020. doi:10.21203/rs.3.rs-66890/v1

116. Williams FN, Chrisco L, Nizamani R, et al. COVID-19 related admissions to a regional burn center: The impact of shelter-in-place mandate. Burns Open 2020;4:158–9. doi:10.1016/j.burnso.2020.07.004

117. Hackett K, Proulx K, Alvarez A. Case Studies of Nurturing Care Initiatives during COVID-19 pandemic.

